# Is there a link between temperatures and COVID-19 contagions? Evidence from Italy

**DOI:** 10.1101/2020.05.13.20101261

**Authors:** Vicente Rios, Lisa Gianmoena

## Abstract

This study analyzes the link between temperatures and COVID-19 contagions in a sample of Italian regions during the period ranging from February 24 to April 15. To that end, Bayesian Model Averaging techniques are used to analyze the relevance of the temperatures together with a set of additional climate, environmental, demographic, social and policy factors. The robustness of individual covariates is measured through posterior inclusion probabilities. The empirical analysis provides conclusive evidence on the role played by the temperatures given that it appears as the most relevant determinant of contagions. This finding is robust to (i) the prior distribution elicitation, (ii) the procedure to assign weights to the regressors, (iii) the presence of measurement errors in official data due to under-reporting, (iv) the employment of different metrics of temperatures or (v) the inclusion of additional correlates. In a second step, relative importance metrics that perform an accurate partitioning of the *R*^2^ of the model are calculated. The results of this approach support the evidence of the model averaging analysis, given that temperature is the top driver explaining 45% of regional contagion disparities. The set of policy-related factors appear in a second level of importance, whereas factors related to the degree of social connectedness or the demographic characteristics are less relevant.

## 1 Introduction

The Coronavirus disease 2019 (COVID-19) pandemic started on December 1, 2019 in Wuhan City, central China, when a group of people with unknown cause pneumonia were reported, mainly linked to workers in the Wuhan South China Wholesale Seafood Market (WHO, 2020a). After the epidemic outbreak in China, on 30 January 2020, the World Health Organization (WHO) Director-General Dr. Tedros declared the outbreak of the novel Severe Acute Respiratory Syndrome Corona Virus (SARS-CoV-2) and the COVID-19 disease “a public health emergency of international concern”. The reason is that the SARS-CoV-2 is a new pathogen that attacks the respiratory system and can lead to an acute respiratory failure or death (Bedford *et al*., 2020; Phua *et al*., 2020; Russell *et al*., 2020).

Despite travel restrictions, border controls, and quarantine measures in China, the epidemic spread worldwide. On 11 March 2020, the WHO declared the COVID-19 “a pandemic disease” after affecting 118,610 people over 114 countries in a matter of weeks. Five weeks later, by April 15, more than 2 million people were infected in 185 countries. The size and strength of the epidemic outbreak has threatened the ability of many health-care systems all over the world to cope with a shock of unprecedented magnitude and produced a severe health crisis. In addition, although the economic impacts of the COVID crisis are yet uncertain, IMF forecasts projects that the “Great Lockdown” employed by governments to contain the disease in many countries is likely to cause a deeper recession than that of the 2008–2009 financial crisis, as the global economy is expected to contract sharply by 3 percent in 2020 (IMF, 2020).

Given that governments all over the world are highly concerned with the threat of the COVID-19, increasing our understanding on the drivers of the geographical variation in COVID-19 contagions is of major importance from a policy perspective. However, to date, little is known about the determinants of regional COVID-19 contagion differentials. From an empirical perspective, the strand of literature analyzing its determinants has highlighted the relevance of different factors in shaping regional reactions to COVID-19 epidemic, such as air pollution (AQR, 2020a; Pansini and Fonaca, 2020; Setti *et al*., 2020; Wu *et al*., 2020), social mobility and connectedness (Arenas *et al*., 2020; Pluchino *et al*., 2020;Kuchler *et al*., 2020), population density (Fang and Wahba, 2020; AQR, 2020b), the level of policy stringency and the timings of the lockdowns (Alvarez *et al*., 2020; Orea and Alvarez, 2020; Casares and Khan, 2020), or the effects of proactive testing (Wang *et al*., 2020b; Romer, 2020) among others.

In this regard, one of the issues that has received more attention is the possible link between climatic factors and contagions (Yao *et al*., 2020; Bukhari and Jameel, 2020; Sajadi *et al*., 2020; Ma *et al*., 2020; Oliveiros *et al*., 2020; Wang *et al*., 2020c) as there is the hope that rising temperatures and humidity during the summer season could reduce SARS-CoV-2 transmission rate, providing time for healthcare system recovery, drug and vaccine development, and a return to economic activity. However, the results are not conclusive as they virtually fit all possibilities and reach diverging conclusions. Yao *et al*. (2020) and Bukhari and Jameel (2020) find no evidence of any statistical relationship between temperatures and contagions, Ma *et al*. (2020), Oliveiros *et al*. (2020), Sajadi *et al*. (2020) and Wang *et al*. (2020c) observe a negative effect, whereas in Merow and Urban (2020) and Pedrosa (2020) a positive relationship is obtained.

These studies represent a substantial progress in understanding the link between climate and the spread of COVID-19 but an important methodological problem present in most of them, is that they have employed very limited sets of variables to analyze this phenomenon and ignored uncertainty surrounding the true model or data generating process (DGP) underlying COVID-19 contagions. From an econometric perspective, ignoring model uncertainty and omitting relevant explanatory variables that could affect COVID-19 patterns, is of major importance given that such estimates may be unreliable (Moral-Benito, 2015, Steel, 2017). Moreover, since it is often not clear a priori which set of variables are part of the “true” regression model, a naive approach that ignores specification and data uncertainty may result in biased estimates, overconfident (too narrow) standard errors and misleading inference and predictions. A second issue with existing analysis on COVID-19 is that they fail to derive a rank of the various factors in terms of their importance, thus, hampering the consensus on what policies could be implemented to increase resilience against this pandemic disease. Third, there is evidence on large cross-country differentials regarding the percentage of detection of COVID-19 cases achieved by each country, which makes difficult to compare existing cross-country incidence series (Russell *et al*., 2020). These measurement errors are likely to plague and contaminate any cross-country analysis and might be aggravated if the ability to test relative to the actual contagions is time-varying, or if there is a varying degree of under-reporting in the number of deaths due to the disease.

To solve the aforementioned problems this study contributes to the literature investigating the linkages between climate and COVID-19 contagions in three major aspects.

First, we investigate the effects of temperatures by means of Model Averaging techniques. This approach allows us to produce a probabilistic ranking of importance through the computation of posterior inclusion probabilities (PIP) for the different variables.^1^ The set of potential determinants considered at the regional level includes: (i) institutional factors and variables related to the regional authorities’ policy response to the epidemic, (ii) demographic factors, (iii) climatic, geographic and environmental characteristics and (iv) variables that capture the degree of social connectedness and mobility of the population. Thus, compared with the limited set of regressors analyzed in the existing empirical literature, this study assesses model uncertainty over a larger set of COVID-19 potential determinants while minimizing omitted variable bias and the multicollinearity concerns that could arise in a single model regression empirical framework. Hence, unlike previous studies using a small fraction of the information available in the data set to draw conclusions, we explore the model space formed by all the possible model combinations implied by the set of potential drivers. In addition, a Posterior Jointness analysis following Doppelhofer and Weeks (2009) is carried out with the aim of increasing our understanding on the factors that might behave as complementary or substitutes to the climate and policy effects on COVID-19 contagions.

Second, to complement the BMA analysis, in a second phase, we calculate relative importance metrics for regression models which allow us to consider all possible causal patterns and channels among the explanatory factors (Johnson and LeBreton, 2004, Gromping, 2007). These metrics perform a decomposition of the *R*^2^ of the regression model, enabling a detailed analysis of the relative contribution of each variable to the explained variability of regional contagion differentials. The main advantage of this approach is that it allows to disentangle the importance of each factor taking into account multivariate interactions with the other factors.

A third point is that instead of employing a non-homogeneous large sample, with observations that are difficult to compare (which usually comes with the additional cost of having a limited set of explanatory factors to investigate regional differentials), here we focus our attention in a small sample of 21-NUTS-2 Italian regions. This research design has some advantages with respect a “large N” design. The first one is that the Italian Civil Protection Ministry (CPM) provides both, longer time-series but also more consistent, reliable and homogeneous regional data in the magnitudes of the health crisis (i.e, number of contagions, deaths, tests, etc) than other advanced countries.^2^ Second, the fact that Italy was one of the countries that was hard hit in the first place by the COVID-19 pandemic, implies that by the time of performing this analysis, we can analyze the full evolution of the first wave of the epidemic, given that abrupt changes in the data are not likely to occur in the near future which could affect the conclusions. Third, by exploiting regional variation within a single country, we minimize the potential biases and problems of data comparability due to under-reporting differentials across countries and across time in both the incidence and death data (see, Russell *et al*., 2020 for a discussion). Nonetheless, as the issue of under-reporting of cases is likely to create measurement errors even in relatively homogeneous sample, we verify if the results obtained are robust with respect to this potential problem.^3^

The study is organized as follows. Section 2, which follows this introduction, reviews the theoretical mechanisms and the empirical evidence linking climate to COVID-19 contagions. Section 3 describes the data on regional contagion and climatic factors and provides preliminary evidence on the link between these variables. Section 4 describes the data set used in this study and the various factors considered in the analysis. In Section 5, the BMA econometric modeling framework is presented. The empirical findings and robustness checks are presented in Section 6, while Section 7 discusses the policy implications than can be derived from this research and offers the main conclusions of the study.

## 2 Why Should Climate Matter for COVID-19 Contagions?

The linkages between viral disease transmission and climatic factors have been well documented in a large number of cases (NCR, 2001). Disease agents and their vectors have specific environments that are optimal for growth, survival, transport, and dissemination. Factors such as precipitation, temperature, humidity, and ultraviolet radiation intensity are part of that environment.

Indeed, each of these climatic factors can have different impacts on the epidemiology of various infectious diseases. For instance, in some viral diseases such as the Dengue or the Malaria, higher temperatures favor spread of the disease whereas in the case of the case of the Influenza higher temperatures and humidity decrease the efficiency of the viral transmission (Lowen and Steel, 2014). In the case of the Influenza, which is transmitted person to person in aerosol droplets typically through coughing and sneezing, the transmission is more efficient at cold temperatures and dry conditions. The specific mechanisms through which cold temperatures increase the diffusion of Influenza are related to the susceptibility conditions of the host or the physical properties of the virion envelope, among others.

In this regard, laboratory studies analyzing both the temperature and humidity effects on the viability of the SARS-CoV, which belongs to the family of the SARS-CoV-2, show that higher temperatures and humidity decrease its viability, as it occurs with the Influenza (Chan *et al*., 2011). In the case of SARS-CoV-2, which is also assumed to be propagated by droplets (WHO, 2020b), two main types of human to human transmission are likely to be at work. The first hypothesis suggests that viral particles emitted from the respiratory tract of an infected individual may land on a surface, another person could touch that object and then touch their nose, mouth or eyes, facilitating the virus to sneak into the body, thereby infecting the second person (WHO, 2020b). The second is that of aerosol transmission. Aerosol transmission is biologically plausible when infectious aerosols are generated by or from an infectious person, the pathogen remains viable in the environment for some period of time, and the target tissues in which the pathogen initiates infection are accessible to the aerosol. Thus, it could be possible that people may emit virus particles in a range of sizes, some of them small enough to be considered aerosols, which can stay suspended in the air for hours and then be inhalated by others (Van Doremalen *et al*., 2020).

The efficiency of these transmission processes is likely be mediated by climatic factors (Duan *et al*., 2003; Lowen and Steel, 2014). A first mediation mechanism is that of temperature and heat. The reason is that colder environments may increase viral transmission given that lower temperatures decrease metabolic functions and mucus secretion, which act as a defense barrier in hosts. Hair-like organelles outside of cells that line the body’s airways, called cilia, do not function as well in dry and cold conditions and they cannot expel viral particles as well as they otherwise would. Secondly, temperatures can affect the transmission through the vehicle itself, the respiratory droplet, as the length of a time a droplet remains airborne depends on its size (Lowen and Steel, 2014). In the winter season, when cold, dry air comes indoors and is warmed, the relative humidity indoors drops sharply, which makes it easier for airborne viral particles to travel. The intuition here is that heating systems increase indoor temperatures and dry-out the air, favoring the evaporation of droplets. In such circumstances, the stronger the evaporation, the smaller the diameter of the droplet, thereby enhancing the airborne suspension and the possibility of aerosol transmission in closed spaces.

A third mechanism linking climate factors to contagions refers to the impacts exerted by sunshine light and ultra-violet (UV) radiation (Merow and Urban, 2020). This is because of there is evidence suggesting that UV radiation might increase human defenses and could reduce virus stability. Sunlight contains different types of UV which might degrade the genetic material of viral particles. Indeed, as shown by Duan *et al*. (2003) UV radiation effectively kills viruses such as the SARS-CoV. Likewise, sunshine light and UV radiation increase vitamin D in humans, which enhances their immune system reducing the likelihood of experiencing respiratory tract infection (Canell *et al*., 2006; Martineau *et al*., 2017).

Taken together, the theoretical arguments outlined above suggest that in regions with higher temperatures, higher humidity and more abundant fluxes of sunshine light or UV, the efficiency of viral transmission could have been lower. However, at the empirical level, it is yet unknown if COVID-19 propagates more slowly in these environments.

Table (1), summarizes the evidence provided by the empirical literature analyzing climate effects on COVID-19. Although there seems that the number of studies finding a negative sign in temperatures and humidity dominate those that find either non-significant or positive effects, the empirical research on the relationship between these variables and COVID has been so far limited and generally reaches diverging conclusions.

The synthesis provided in Table (1) shows that there are at least four studies finding that temperatures reduce COVID-19 transmission, two of them observe that the range of temperatures does not matter and the other two obtain that temperatures could in fact have a positive impact on contagions. In the case of UV there are just two studies, one with a non significant link and the other one with negative effect, whereas in the case of humidity, three of them report a reduction effect on COVID viral transmission and the other three a positive one.

**Table 1:**
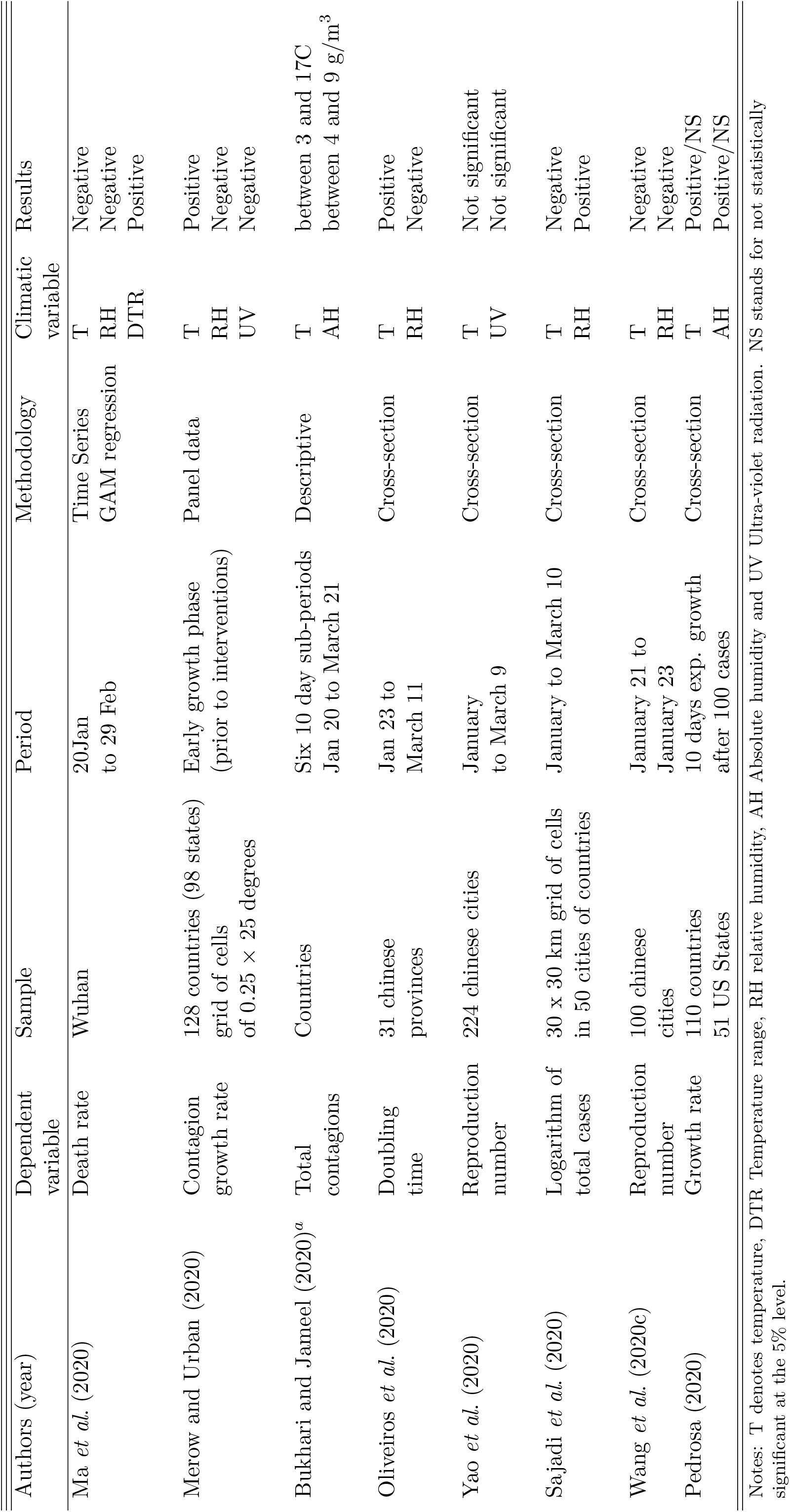
The Empirical Relationship between Climate and COVID.

Specifically, Ma *et al*. (2020) investigate the effect of temperatures and humidity on deaths in Wuhan by means of a time-series Generalized Additive Model (GAM). By exploiting time variation in temperature data of Wuhan from the 20 of January to the 29 of February, and controlling for air pollutants, they find a negative and statistically significant effect of ambient temperatures and humidity. Sajadi *et al*. (2020) investigate the link between climate factors within a sample of 50 cities of different countries all over the world by means of linear regression techniques finding that temperatures 2 meters above the surface, exert a negative and statistically significant effect on the total number of contagions whereas humidity increases them.

Other two studies focusing on cross-sectional variation among chinese cities find a negative and statistically significant link between temperatures and contagion speed. Oliveiros *et al*. (2020) use data from 31 chinese provinces and linear regression modeling. In their study they observe a negative link between median temperatures after controlling for humidity, wind speed and precipitation. On the other hand, Wang *et al*. (2020c) observe a negative link between temperatures and the reproduction number of the COVID19 in 100 chinese cities for the pre-intervention period, after controlling for relative humidity, GDP per capita, population density, the number of hospital beds and the share of population over 65. The effect of humidity in this analysis appears to be detrimental to contagions.

As opposed to these two papers, Yao *et al*. (2020) within in a sample of 232 Chinese cities and after adjusting for relative humidity and UV, report that temperatures held no significant associations with cumulative incidence rate.

The descriptive analysis of Bukhari and Jameel (2020) finds that 90% of contagions occurred in a wide range of temperatures and humidity, between 3 to 17 Celsius degrees and suggests that reduced spreading due to environmental factors would be limited in most of northern Europe and North America in the summer season. Merow and Urban (2020) using a panel data of 128 countries including country-fixed effects and daily data on contagions obtain a positive effect of the 14 day lag of mean temperatures on the growth rate of contagions, once the proportion of elderly, relative humidity and ultraviolet light are taken into account. Similarly, Pedrosa (2020) finds a non robust positive effect of temperatures in a sample of US states and in a sample of 110 countries by looking at the early growth rate of the contagions.

The reasons for this diversity of results have to do with the fact that these contributions differ considerably in terms of the sample composition, the study period, the indicator used to measure the impact of the epidemic and the temperatures, and more importantly, in the econometric and statistical approach employed to perform inference. Moreover, as discussed in Section (1) a drawback of existing literature is that it has focused on a limited number of variables and explanatory factors, which casts doubts on the validity of these findings (i.e, Yao *et al*. (2020) or Sajadi *et al*. (2020) do not include any covariate). Accordingly, further empirical research is required to clarify the nature of the link between climatic factors and COVID-19 incidence at the regional level.

## 3 The Link between COVID-19 Contagions and Temperatures in Italian Regions

This section provides preliminary evidence on the linkages between climate and COVID-19 incidences in Italian regions.

To that end, we first describe the official data on incidences at the regional level in Italy, up to April 15. Data were collected daily at 12 p.m. (GMT+2), between February 24, 2020 to April 16, 2020 from the Italian Ministry of Civil Protection (MCP).

The analysis considers NUTS-2 level regions rather than other possible alternatives for various reasons. Firstly, the use of data at the NUTS-2 aggregation level allows us to built an homogeneous database on a larger number of potential drivers of contagions using distinct sources. Secondly, NUTS-2 is the territorial unit most commonly employed in the literature on regional analysis and it is particularly relevant in terms of Italian health-care regional policy, given that health competences in Italy are decentralized (Golinelli *et al*., 2017).^4^

However, before continuing, it is necessary to stress that the quality of the available contagion data on COVID-19 is far from being optimal since the time-series on infections are affected by a severe share of under-reported cases and Italy is no exception in this regard (see, Russell *et al*., 2020). The reason is that only the more severe cases (non-asymptomatic and non-mild) cases get sick enough to search medical help and have been officially diagnosed. Therefore, the quality of the data on contagions depends to a large extent on the fraction of cases that are severe enough to both lead to medical attention and be tested. Hence, by using this metric on contagions, we are measuring “two phenomena at the same time”: the infected people and the authorities’ ability to detect them. Note that although this point could be a major drawback in cross-country comparative research, it is reasonable to assume that within a country, these measurement errors do not differ significantly across regions, and as a consequence, these concerns are of less importance in this context.^5^

To characterize the dynamics of COVID-19 incidence at the regional level, we begin by analyzing changes in the evolution of the official time series of reported contagions per capita. Figure (1) shows the evolution of cumulative contagions per 100,000 inhabitants from February 24, 2020 to April 15, 2020. As observed, the figures increased substantially during March 2020, such that by March 21, all the regions in the country had more than 10 COVID-19 contagions per 100,000 inhabitants.

**Figure 1:**
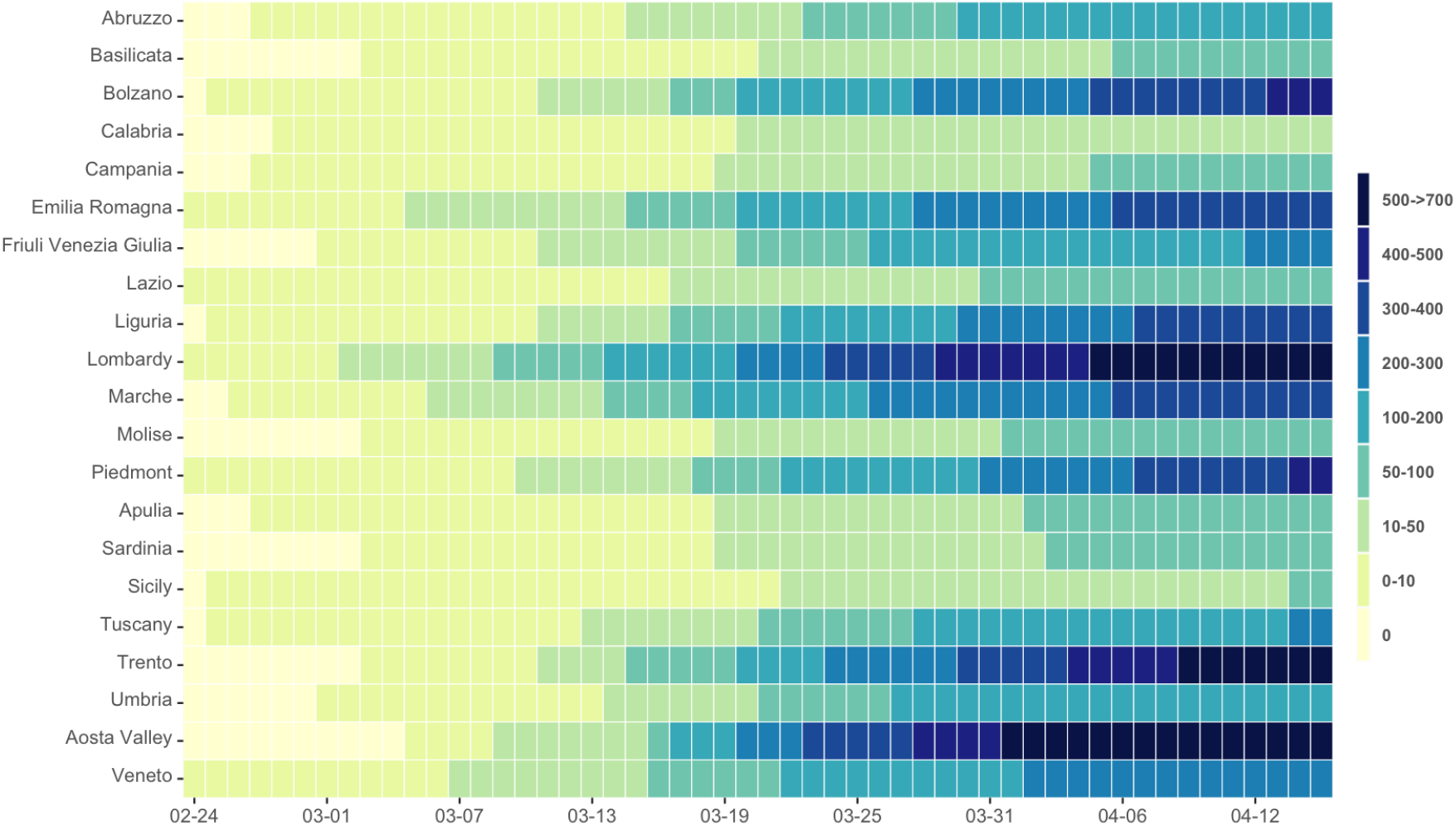
The dynamics of Contagions in Italian regions

As shown in Figure (1), the region that increased more rapidly the number of infections and crossed the 10 cases per 100,000 inhabitants threshold in a first place was Lombardy, on March 2. By March 24, Lombardy had more than 300 cases and from April 5 onwards, the figures in Lombardy located in the interval ranging from the 500 to 700 cases per 100,000 inhabitants. The evolution of Lombardy was closely followed by the regions of Marche, Emilia-Romagna, Bolzano and Aosta Valley, who also experienced markedly increases, and before March 22, all the neighbors of Lombardy located in the north of the country reported more than 100 cases per 100,000 inhabitants.^6^ This pattern of contagions confirms a rapid cross-regional diffusion such that most of the Italian regions arrived to their peak of contagions per capita during the last ten days of March and stabilized their growth rates during the first half of April.^7^

To carry out our research and explore the effects of temperatures we also need climate data. Given that there is a time lag between person to person transmission, the development of symptoms, the test realization and the confirmation of test results later on, we resort to a lagged value of temperatures to capture the fact that if present, climatic effects should have taken some time to exert an impact on contagions. For this reason, we employ the average temperatures in February of each regional centroid, obtained after averaging over the time series of daily mean temperatures at 2 meters above the surface of the earth. The meteorological data was taken from the NASA-Prediction Of Worldwide Energy Resources (NASA-POWER) v8 GIS database.

Figure (2) panel (a) shows the geographical distribution of the total contagions per 100,000 inhabitants for each region in April 15. An interesting fact shown in Figure (2) is that the geographical distribution of infections has been clearly asymmetric across regions. On the other hand, the information of temperatures is shown in the panel (b) of Figure (2). As the color scale range in each panel is set to reflect a higher value of the variable being mapped, the observation of Figure (2) reveals that higher cumulative incidence levels up to April 15, occurred in regions with lower temperatures in February.^8^

**Figure 2:**
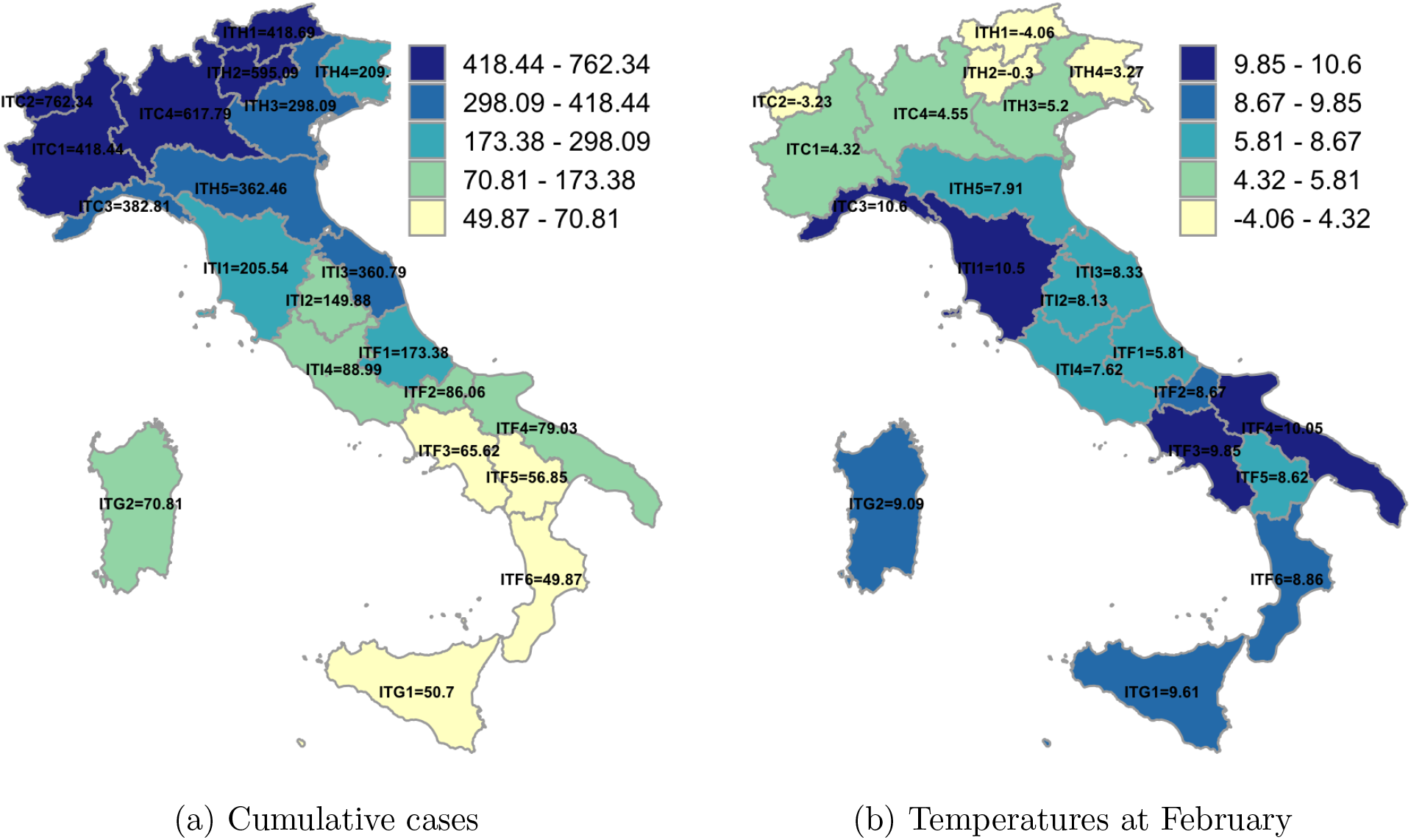
The geographical distribution of contagions and temperatures

To further investigate this link, Figure (3) provides a graphical illustration on the association between average daily temperatures and COVID-19 cumulative incidences per 100,000 inhabitants by April 15. The scatter plots suggests the existence of a negative relationship between the magnitude of cumulative contagions and temperatures. This means that regions with higher temperatures tend to have lower levels of infections, while those regions with lower temperatures, on average, are characterized by a higher incidence. Indeed, the pairwise correlation between the two variables is statistically significant (*ρ* = −0.723 with p-value = 0.00).^9^. Nonetheless, the information provided by Figure (3) should be treated with caution, as the observed connection between temperatures and the regional epidemic size may simply be a spurious correlation resulting from an ecological fallacy or the omission of other variables affecting both, climate and the infections. In view of this potential problem, in Section (5) we develop a more appropriate statistical analysis on the link between temperatures and regional COVID-19 cases.

**Figure 3:**
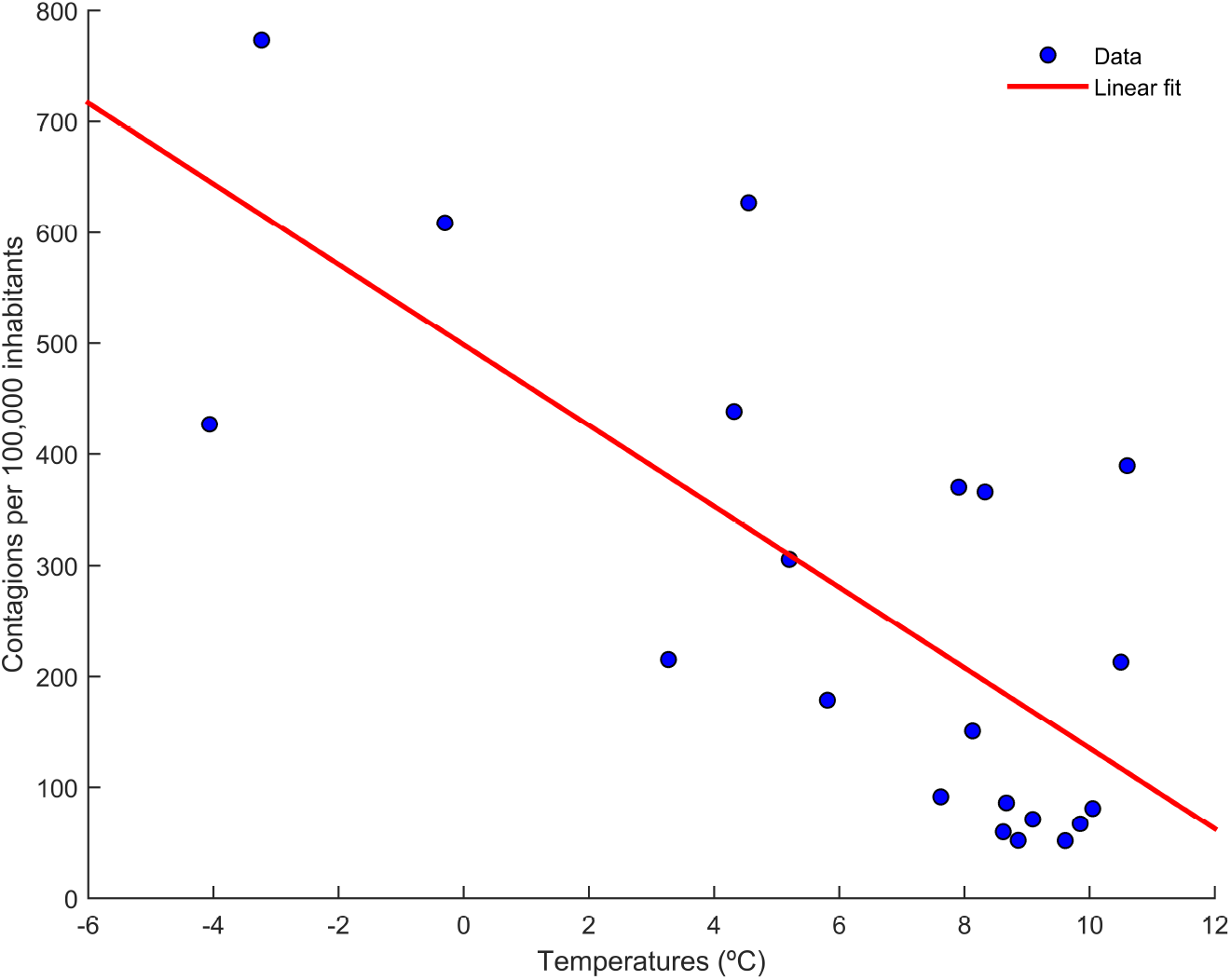
The link between Temperatures and COVID-19 Contagions in Italian Regions

## 4 The Determinants of COVID-19 Contagions

To produce a comprehensive ranking on the importance of the diverse potential factors influencing contagion differentials during the epidemic outbreak, we consider a variety of candidate factors. Specifically, we consider (i) institutional characteristics and variables related to the policy response during the epidemic, (ii) demographic characteristics, (iii) climate and environmental characteristics and (iv) the degree of social connectedness among the population. We now describe these controls and provide a brief conceptual justification for their inclusion in the analysis. Table (A2) in the Appendix presents the detailed definition and sources of all the control variables used in the paper.

### 4.1 Institutions and Policy

The first variable considered is (i) the ability of regional authorities to perform tests and detect infections in an early stage. We expect this variable to have decreased contagions given that an early and correct diagnose of the infection may have helped to identify potential carriers, isolate them properly and reduce the spread of the epidemic (Romer, 2020; Wang *et al*., 2020b).^10^ To proxy the ability of monitoring the epidemic in advance we use the number of tests per 100,000 inhabitants during the first week of the epidemic outbreak, from the 24 of February to March 1.

A second factor included refers to the (ii) time delay or policy lag in the application of regional lockdowns to limit viral transmission. Given that during the early stages of the COVID-19 outbreak the number of new cases was growing at an exponential rate, time lags in the adoption of the regional lockdown of territories relative to the stage of the epidemic outbreak may have helped to increase diffusion and total pandemic size (Alvarez *et al*., 2020; Orea and Alvarez, 2020; Casares and Khan, 2020). In Italy, early lockdowns were adopted by March 8 in the regions of Lombardy, Marche, Emilia Romagna, Veneto and Pidemont covering 16.46 million inhabitants. Two days later, a nation-wide lockdown was extended to all the regions and the whole population. We proxy the lag in the policy response by calculating the number of days with more than 1 case per 100,000 inhabitants prior to the lockdown of the region.

Finally, we consider (iii) the ratio of health expenditures to GDP, because of regions with a higher expenditure are more likely to have experienced less shortages of equipment and educated manpower to implement tests, carry out treatments and ultimately, minimize within-hospital contagion amplification, also known as “nosocomial transmission” (Bedford *et al*., 2020). In addition, regions with a higher share of resources of their GDP devoted to health-care services may have been able to perform intensified contact tracing and locate cases to stop onward transmission. Thus, we expect a negative effect of higher health expenditures.

### 4.2 Demographic factors

Socio-demographic factors may also play a relevant role determining reported contagions per capita. In particular, the age composition of the population is expected to have an impact on contagions. To control for differences in the demographic structure we consider (iv) the share of population under 35 years old and (v) the share of population above 75 years old. Given that there is evidence that effects of the COVID-19 are much more severe in older populations (Verity et al., 2020) and because of the limited ability of performing tests on the population, regional authorities may have prioritized detection among the elderly with respect the younger (which in many cases could have been asymptomatic carriers). For this reason we expect regions with a higher share of elderly population to have reported more cases per capita than regions with a younger population.

### 4.3 Social connectivity factors

We also consider the fact that different degrees of social connectivity could also have an impact in the spread of the disease and the probability of importing infections from abroad (Charaudeau *et al*., 2014). To control for regional differentials in this regard, we first consider (vi) the degree of social mobility of the population, which is expected to increase contagions (Arenas *et al*., 2020; Pluchino *et al*., 2020;Kuchler *et al*., 2020). Following Pluchino *et al*. (2020) we measure the mobility of the population as the ratio between the sum of commuting flows (incoming and outgoing) for each municipality in a region and the population employed in that municipality using data from the Italian Ministry of Economic Policy Planning and Coordination. The regional value is obtained by averaging over all municipalities within the region.

A second candidate factor to explain these differentials is the (vii) population density, because of the transmission of SARS-CoV-2 through the population operates through close person-to-person contact, most readily by respiratory droplets, whether through polluted surface contact (WHO, 2020b) or aerosol (Van Doremalen *et al*., 2020). Thus, compared to low-density areas, a higher population density could favor the interaction and personal contacts reinforcing the transmission of the virus (AQR, 2020b). Nevertheless, this argument should be taken with caution, as some extremely dense cities, such as Singapore, Seoul, and Shanghai, have outperformed many other less-populated places in combating the coronavirus. As a point of fact, Fang and Wahba (2020) find no statistical relation between density and contagions in a sample of Chinese cities.

### 4.4 Climate and environmental factors

The third group of factors that are likely to explain contagions differentials refer to climate and the environmental conditions of the region. Air pollution is considered a factor that may exacerbate the vulnerability of populations by worsening chronic lung and heart infections (Cui *et al*., 2003). In the context of the COVID-19 existing studies have failed to stablish a clear causal link between particulate pollution, contagion and the subsequent health damage (Pansini and Fonaca, 2020; Wu *et al*., 2020; Wang *et al*., 2020a). However, the intuition for a positive link is that airborne particles may have been able to serve as carrier for the pathogen (Setti *et al*., 2020). Whether the virus remains viable after hitching a ride on pollution particles, or whether it can do so in sufficient amounts to cause infection is unclear. In any case, to control for this potential determinant, we account for (viii) environmental quality differentials. We measure the impact of environmental quality by means of the share of population that is exposed to more than 15 micrograms/m3 of particulate matter (PM2.5) using data from the OECD. Our expectation is that a higher pollution may have exerted a positive effect on the pandemic size.

Makinen *et al*. (2009) find that in general, when the average temperature drops the estimated risk for the lower respiratory tract infections increases. Apart of the potential role of (ix) temperatures, another climatic factor that has received considerable attention in the literature is that of (ix) relative humidity (Oliveiros *et al*., 2020; Sajadi *et al*., 2020, Wang *et al*., 2020c). As discussed in Section (2), we expect regions with lower temperatures and lower humidity to have a higher incidence as these two factors may have reduced the efficiency of viral transmission by decreasing host resistance in outdoor environments and increasing the stability of airborne viral particles in indoor spaces.

## 5 Econometric Strategy: Bayesian Model Averaging

In empirical COVID-19 research, although some variations of a baseline model are often reported, basing inference on a single model has become a common practice. Typically, researchers draw their conclusions on this single model acting as if the model was the true model. Nevertheless, this procedure uses a minimal fraction of the information in the data set and understates real uncertainty associated with the specification of the empirical model (see, Moral-Benito, 2015; Steel, 2017). To address this concern, in our analysis of the drivers of COVID-19 regional differentials we employ the Bayesian Model Averaging approach. We begin by considering the following regression model:

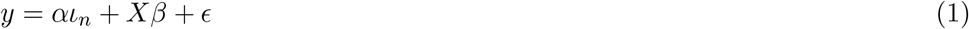

where *y* denotes a *N* × 1 dimensional vector consisting of observations for the cumulative total infections per capita during February 24 to April 15, for each region *i* = 1*, …, N*. *α* reflects the constant term, *ι_n_* is a *N* × 1 vector of ones, *X* is an *N × K* matrix of regional explanatory variables with associated response parameters *β* contained in a *K* × 1 vector. Finally, ∈ = (*ϵ*_1_*, …, ϵ_N_*)′ is a vector of i.i.d disturbances whose elements have zero mean and finite variance *σ*^2^.

Bayesian Model Averaging (BMA) techniques haven been previously analyzed to investigate the determinants of complex processes in the field of regional science (Crespo-Cuaresma *et al*., 2014, Hortas-Rico and Rios, 2019, Rios and Gianmoena, 2020) and a large literature on BMA in regression models already exists (see Moral-Benito, 2015, Steel, 2017) for detailed reviews on the literature). However, to get an intuition behind the BMA approach employed here to learn on the nature of COVID-19 regional disparities, notice that for any set of possible explanatory variables of size *K*, the total number of possible models is 2^*K*^ and *k* 0, 2^*K*^. This implies there are 2^*K*^ sub-structures of the model in Equation (1) given by subsets of coefficients *η^k^* = *α, β^k^* and combinations of regressors *X_k_*.^11^ Model averaging techniques solves the question of variable importance and the effects of each regressor *h* by estimating all the candidate models implied by the combinations of regressors in *X* (or a relevant sample of them) and computing a weighted average of all the estimates of the corresponding parameter of *X_h_* (where the sub-index *h* here denotes a single regressor and not a model or a combination of regressors *k*). By proceeding in this way, estimates consider both the uncertainty associated to the parameter estimate conditional on a given model, but also the uncertainty of the parameter estimate across different models.

By following the Bayesian logic, the posterior for the parameters *η_k_* calculated using model *M_k_* is written as:

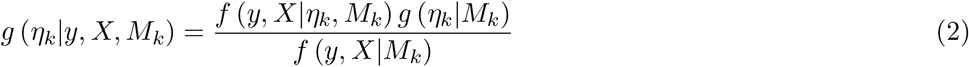

where *g* (*η_k_*|*y, X, M_k_*) is the posterior, *f* (*y, X*|*η_k_, M_k_*) is the likelihood and *g* (*η_k_*|*M_k_*) is the prior. The key metrics in BMA analysis are the Posterior Mean (PM) of the distribution of *η*:

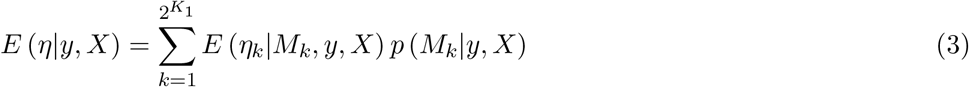

and the Posterior Standard Deviation (PSD):

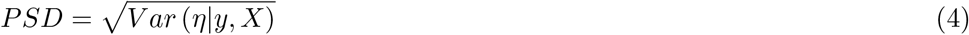

where the *V ar* (*η*|*y, X*) is given by:

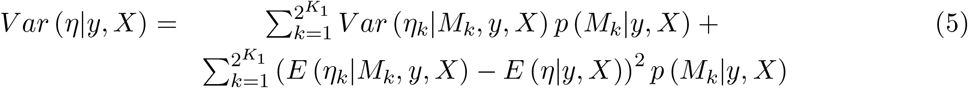

To derive these metrics, it is necessary to calculate the Posterior Model Probability *p* (*M_k_*|*y, X*) of each of the sub-models *M_k_*. These can be obtained as:

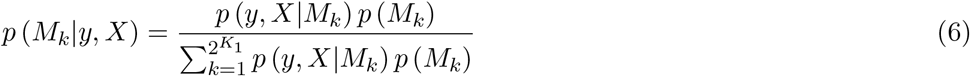

where *p* (*y, X*|*M_k_*) is the marginal likelihood and *p* (*M_k_*) is the prior model probability. The marginal likelihood of a model *k* is calculated as:

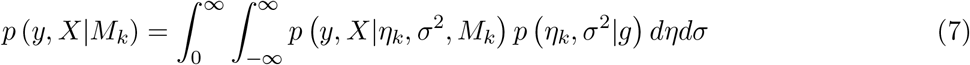

where *p y, X*|*η_k_, σ*^2^*, M_k_* is the likelihood of model *k* and *p η_k_, σ*^2^|*g* is the prior distribution of the parameters in model *M_k_* conditional to *g*, the Zellgner’s g-prior.

In addition, the BMA framework can be extended to generate probabilistic on the relevance of the various regressors, using the Posterior Inclusion Probability (PIP) for a variable *h*:

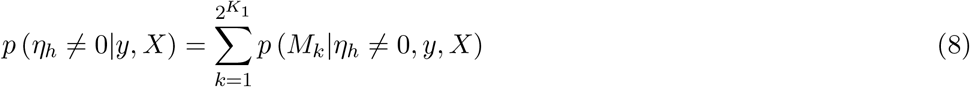

and the Conditional Posterior Positivity of *h*:

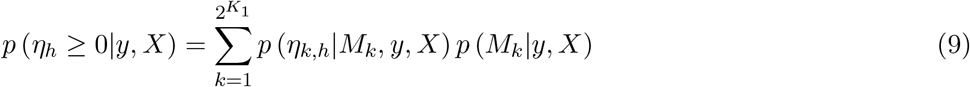

where values of conditional positivity close to 1 indicate that the parameter is positive in the vast majority of considered models and values close to 0 indicate the effect on the dependent variable is negative.

The calculation of previous metrics in the BMA approach requires to define priors on the model space and priors on the parameter space. We use a model specific Zellgner g-prior based on the Bayesian Risk Inflation Criterion (BRIC), whereas we use a calibrated Binomial prior on the model space to give the same a priori probability to each model and variable.^12^

Following early discussion of Bayesian measures of variable importance, PIPs in Equation (8) have become a standard tool for interpreting the results in econometric applications of BMA (Doppelhofer and Weeks, 2009). However, although they provide valuable insight into the overall importance of single variable, they neglect the interdependence of inclusion and exclusion of variables. Thus, PIPs do not help to conclude if the importance of the variable is evenly spread out across all model specifications or if it is specific to a certain combination of explanatory variables. To gain insights into the interdependence of the inclusion of sets of different variables, several studies investigate the joint posterior of pairs of variables. Jointness reveals generally unknown forms of dependence. Positive jointness implies that regressors are complements, representing distinct but mutually reinforcing effects. On the other hand, negative jointness implies that explanatory variables are substitutes and capture similar underlying effects. To analyze this issue, we follow Doppelhofer and Weeks (2009), who propose the use of log of a cross-product ratio of inclusion probabilities. For any pair of arbitrary variables A and B of the set of *K* potential variables, we calculate their bivariate jointness as:

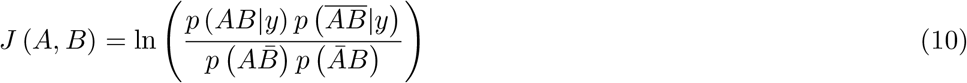

where values of the *J* statistic above 1 are considered as evidence on significant complementarity, values below –1 suggest significant substitutability and values between −1 and 1 are suggest independence. 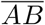 indicates that event AB did not occur, 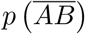 denotes the probability of models where AB did not occur, etc.

## 6 Results

### 6.1 Main BMA Results

Table (2) reports the results obtained from the BMA analysis. However, before continuing with the discussion of the results, it is worth mentioning the problems that the methodology applied here can solve and those problems that may persist, affecting the quality of the estimates. The strong point of the BMA methodology employed here is that it accounts for the uncertainty of the parameter estimates across different models while controlling for omitted variable bias and reducing multicollinearity problems (Moral-Benito, 2015; Steel, 2017). Nevertheless, it does not correct for the potential negative effect of endogeneity generated by reverse causal relationships, or measurement errors. The issue of reverse causal relationships is likely to be at work in the case of the proactive testing proxy variable, whereas measurement errors can be an issue in the dependent variable, the number of contagions per 100,000 inhabitants. Therefore, to minimize the potential problems caused by reverse causality relationships and the lack of an appropriate instrument, tests are measured with respect the total population during the first week of the epidemic (i.e, 6 weeks before the measurement of the dependent variable). On the other hand, the issue of measurement errors in the number of contagions is addressed in detail in Section (6.2.2).

**Table 2:** Main Results: Bayesian Model Averaged Estimates.

| Variable | PIP | Cond. Post.<br>Estimates<br>Estimates | Cond. Post<br>Standardized | Cond. Post.<br>Sign<br>Positivity |
| --- | --- | --- | --- | --- |
|  | (1) | (2) | (3) | (4) |
| Temperatures | 0.979 | -34.296***<br>(7.10) | -0.683***<br>(0.141) | 0.000 |
| Policy Lags | 0.880 | 28.230***<br>(9.01) | 0.509***<br>(0.16) | 1.000 |
| Relative Humidity | 0.440 | -12.872**<br>(6.43) | -0.222**<br>(0.11) | 0.000 |
| Tests | 0.300 | -1.755<br>(1.10) | -0.219<br>(0.14) | 0.007 |
| Health spending | 0.174 | -34.027<br>(33.12) | -0.292<br>(0.284) | 0.001 |
| Share of old population | 0.141 | -8.092<br>(35.03) | -0.059<br>(0.26) | 0.212 |
| Social Mobility | 0.139 | 10.549<br>(17.44) | 0.149<br>(0.25) | 0.749 |
| Share of young population | 0.120 | -9.373<br>(22.78) | -0.090<br>(0.218) | 0.203 |
| Population density | 0.118 | 0.160<br>(0.31) | 0.086<br>(0.17) | 0.954 |
| Air pollution | 0.118 | 0.919<br>(1.52) | 0.078<br>(0.13) | 0.940 |

Notes: The dependent variable in all regressions is the total number of contagions per 100,000 inhabitants from the Italian MCP. The results reported correspond to the estimation of all the possible regressions including any combination of the 10 variables. Prior mean model size is 5. Variables are ranked by Column (1), the posterior inclusion probability. Columns (2) to (3) reflect the conditional posterior mean and the standardized conditional posterior mean for the linear marginal effects of each variable, respectively. Column (4) is the sign certainty probability, a measure of our posterior confidence in the sign of the coefficient. Standard deviations in parenthesis.
\* Significant at 10% level, \*\* significant at 5% level, \*\*\* significant at 1% level.

We use the PIPs of the different variables to classify evidence of robustness of the different contagions drivers, such that regressors with PIPs above the a priori inclusion probability (APIP) are considered as relevant determinants and variables with PIP < APIP, as irrelevant ones. From a Bayesian perspective, variables with PIPs higher than others, reflect a higher importance as they are more likely to be part of the DGP and as such, they can be considered as a relevant piece in explaining contagion differentials across Italian regions.^13^ As observed in Column (1) of Table (2), there is a small group of top variables conforming the group of important determinants. These are the mean temperatures (97.5%) and the policy lags (88%), which are both statistically significant at the 1% level. In the frontier of relevance/irrelevance we also find the variable measuring the relative humidity conditions, with a PIP of the 44%, but with a statistically significant effect on contagions. It is somehow surprising that none of the remaining factors considered, although display the expected signs and are mostly consistent with those obtained in the literature (i.e, a positive link between contagions and the share of old population, social mobility, population density and air pollution; and a negative one between the share of young, health spending and tests), do not appear to be relevant in explaining regional differentials across Italian regions. Overall, our findings suggest, that by focusing only on the role played by climate conditions and the timings of the policy response, we have enough information to explain why some regions may have been affected more than others.

We now turn our attention to the model averaged estimates of our regional-level variables as they provide the basis for posterior inference regarding the parameters. Model averaged estimates of each variable are constructed using all possible model combinations implied by our set of variables and computing a probabilistically weighted average of all the estimates of the corresponding parameter. The results displayed in Table (2) correspond to the estimation of all the models in the model space including any combination of the 10 variables.^14^ The estimated posterior model size has a mean of 3.4 variables, lower than its expected a priori size of 5 variables, which is in line with the finding of very few candidate determinants being relevant and a parsimonious model specification.

As regards our variable of interest, we find that temperatures exert a negative impact on regional contagion outcomes with a posterior mean of –34.296 and a standardized mean effect of –0.683. This means that an increase in 1 Celsius degree in the mean regional temperature during February, decreased the number of total contagions per 100,000 inhabitants in April 15 by 34.29. Alternatively, the estimated standardized posterior mean coefficient in Column (3) reveals that on average, a 1 standard deviation shock to the temperatures (i.e, a change of about 4.3 celsius degrees, which is the difference in the temperatures between the average temperatures of Lombardy (ITC4) and Calabria (ITF6)) had the effect of decreasing the total number of contagions per 100,000 inhabitants by 147.54, which is the difference between the highly affected region of Marche (ITI3) and that of Tuscany (ITI1). Moreover, as shown in Column (4), the posterior sign positivity of the temperatures is 0.00%, which implies the parameter estimate is always negative irrespective of the model in which the variable appears. This negative effect of temperatures on regional contagions is in line with previous literature and supports the findings of Ma *et al*. (2020), Oliveiros *et al*. (2020), Sajadi *et al*. (2020) and Wang *et al*. (2020c) while contradicts those of Merow and Urban (2020), Pedrosa (2020) and Yao *et al*. (2020). As explained, before, the rationale for a negative effect of temperatures on the number of contagions has to do with the increased susceptibility of the hosts due to a slower metabolism decreasing their defenses, and the fact that in winter, when cold air comes indoors and is it warmed by our heating systems, the relative humidity indoors drops sharply facilitating airborne viral particles to travel and be transmitted from one person to another as an aerosol.

We also find the effect of relative humidity helps to explain part of the observed regional heterogeneity. In particular, the estimated effect is negative in all the models, which suggests that regions with higher outdoor humidity experienced lower contagion rates. This finding supports previous results of Wang *et al*. (2020c) and Ma *et al*. (2020).

The finding of a positive and statistically significant impact on the number of infections due to a late political response, is also in line with the results obtained in other studies (Orea and Alvarez, 2020; Alvarez *et al*., 2020). Specifically, we find that each additional day in which the lockdown was not implemented, with respect to the detection date of 1 case per 100,000 inhabitants, meant an increase of 28.230 cases per 100,000 inhabitants by April 15. To put this figure in context, this estimates imply that in Lombardy, implementing the full lockdown in March 1 instead of the 8 of March, would have decreased the total amount of cases by 32% decreasing the incidence toll from 62,153 cases to 42,226 cases. This result is lower than the estimates provided in the empirical study of Orea and Alvarez (2020) who find that for Spanish regions the implementation of the lockdown one week before the date it occurred, had a reduction effect of the cases of approximately the 50%. This difference in the magnitudes can be explained by the fact that even if Italy was one of the first countries implementing the lockdown, in relative terms, the date when this policy measure took place was late, such that the potential of flattening the epidemic curve was lower.

To complement previous results and to gain further insights that might be useful for policy-making, we carry out an analysis of the posterior jointness to detect PIP dependencies among regressors. Table (3) reports the posterior jointness relationships of the different determinants included in the analysis, as calculated by the metric proposed by Doppelhofer and Weeks (2009). The investigation of the jointness is relevant from a policy-making perspective as it helps to better understand what regional factors may reinforce or hamper the efforts of increasing resilience against the COVID-19 epidemic outbreak.

**Table 3:** Posterior Jointness.

|  | Health<br>Spending | Social<br>Mobility | Temp | Pop.<br>dens | Young<br>pop. | Old<br>pop | Tests | Policy<br>Lag | Air<br>Pollution | Rel.<br>Humidity |
| --- | --- | --- | --- | --- | --- | --- | --- | --- | --- | --- |
| Health spending |  | -0.28 | **-4.32** | -0.24 | -0.24 | -0.23 | -0.51 | **-1.49** | -0.35 | -0.50 |
| Social Mobility | -0.28 |  | -0.29 | -0.35 | -0.13 | -0.08 | -0.22 | **-1.43** | -0.18 | 0.22 |
| Temperatures | **-4.32** | -0.29 |  | -0.66 | -0.09 | -0.13 | 0.79 | **2.97** | -0.21 | **1.23** |
| Pop. density | -0.24 | -0.35 | -0.66 |  | 0.21 | 0.03 | -0.29 | -0.89 | -0.32 | -0.29 |
| Young pop. | -0.24 | -0.13 | -0.09 | 0.21 |  | -0.27 | -0.39 | -0.91 | -0.30 | -0.43 |
| Old pop. | -0.23 | -0.08 | -0.13 | 0.03 | -0.27 |  | 0.16 | -0.78 | -0.21 | -0.09 |
| Tests | -0.51 | -0.22 | 0.79 | -0.29 | -0.39 | 0.16 |  | **1.17** | -0.31 | 0.46 |
| Policy Lags | **-1.49** | **-1.43** | **2.97** | -0.89 | -0.91 | -0.78 | **1.17** |  | -0.02 | 0.30 |
| Air Pollution | -0.35 | -0.18 | -0.21 | -0.32 | -0.30 | -0.21 | -0.31 | -0.02 |  | -0.48 |
| Rel Humidity | -0.50 | 0.22 | **1.23** | -0.29 | -0.43 | -0.09 | 0.46 | 0.30 | -0.48 |  |

Notes: the values refer to the J statistic proposed by Doppelhofer and Weeks(2009). The results reported correspond to the estimation of all the models in the model space including any combination of the 10 variables.

The main findings are as follows. We find evidence of both positive and negative jointness among contagion determinants. Significant positive jointness (with J > 1) is not restricted to variables considered significant by the PIPs. In this respect, a more complex explanation emerges, given that variables such as the health spending ratio to the GDP, which seems to decrease contagions, appears to be a strong substitute of policy lags and temperatures. This suggests that in the absence of a warm climate or a fast policy response, a higher health spending to GDP ratio could become a relevant factor to decrease contagions. This can be attributed to both, the higher ability of monitoring and isolating cases and the possibility to reduce “nosocomial transmission” because of an increased access to protection equipment. Other co-dependencies with respect the policy lags are worth mentioning. Conditional to a delayed implementation of the lockdowns, we observe that a pro-active testing policy to detect infections becomes a more relevant determinant and may help to decrease contagions. In addition to this, we find a high degree of substitutability between policy lags and social-mobility, which suggests that restrictions to mobility may produce a similar effect to those of the lockdown of the entire region. Therefore, there results of this posterior jointness analysis reveal that there is room for policy-makers to increase resilience against the epidemic. Place-based oriented policies aiming at increasing health-care capacities and resources, or investments that help workers and firms to adopt teleworking practices that reduce social mobility might be important for the re-activation of the economy while keeping the number of contagions down.

### 6.2 Robustness checks

We now investigate if the finding of temperatures being the most relevant determinant of contagion differentials across Italian regions is robust to changes in the set-up of the analysis. Specifically, in Section (6.2.1) we first analyze if consider the results regarding the probabilistic importance of the variables are being driven by the prior-distributions placed on the parameters and the distribution of the model size. In Section (6.2.2) we check if the rankings are robust to the importance weighting procedure. To that end, we develop a novel frequentist model averaging framework extending Hansen and Racine (2012) which produces importance ranking using jack-knife model averaged variable weights. In Section (6.2.3) we study if correcting for measurement errors in the number of contagions affects the validity of previous findings. In addition, we analyze whether the results are affected by using alternative measurements of temperatures such as the minimum or the maximum temperatures, or if using data from a different source, matters for the validity of our conclusions. Finally, in Section (6.2.4) we check if the rankings are robust to the extension of the model space by increasing the number of candidate determinants.

#### 6.2.1 Prior Distributions

An implication of the Bayesian Model Averaging approach is that inferences drawn on the effects of the various factors and their importance, depend on prior distributions assigned to the model parameters and on the model space. To verify the conclusions do not depend on subjective prior information implied by the elicitation of our prior distributions on the parameters and the model space, we now discuss the results of the different robustness checks performed along this line. Table (4) serves to that purpose.^15^

**Table 4:** Robustness checks (I): The role of priors.

|  | Model Size=5 |  | Model Size=6 |  | Model Size=7 |  | Model Size=8 |  |
| --- | --- | --- | --- | --- | --- | --- | --- | --- |
|  | Prior | Posterior | Prior | Posterior | Prior | Posterior | Prior | Posterior |
| Temperatures mean | 0.500 | 0.979 | 0.600 | 0.989 | 0.700 | 0.993 | 0.800 | 0.996 |
| Policy Lags | 0.500 | 0.880 | 0.600 | 0.906 | 0.700 | 0.925 | 0.800 | 0.949 |
| Relative Humidity | 0.500 | 0.440 | 0.600 | 0.569 | 0.700 | 0.806 | 0.800 | 0.828 |
| Tests | 0.500 | 0.300 | 0.600 | 0.436 | 0.700 | 0.600 | 0.800 | 0.771 |
| Health spending | 0.500 | 0.174 | 0.600 | 0.200 | 0.700 | 0.255 | 0.800 | 0.343 |
| Share of old population | 0.500 | 0.141 | 0.600 | 0.209 | 0.700 | 0.311 | 0.800 | 0.455 |
| Social Mobility | 0.500 | 0.139 | 0.600 | 0.192 | 0.700 | 0.273 | 0.800 | 0.382 |
| Share of young population | 0.500 | 0.120 | 0.600 | 0.163 | 0.700 | 0.231 | 0.800 | 0.336 |
| Population density | 0.500 | 0.118 | 0.600 | 0.165 | 0.700 | 0.230 | 0.800 | 0.328 |
| Air Pollution | 0.500 | 0.118 | 0.600 | 0.165 | 0.700 | 0.235 | 0.800 | 0.344 |
|  | RIC |  | UIP |  | EBL |  | Hyper-g |  |
|  | Prior | Posterior | Prior | Posterior | Prior | Posterior | Prior | Posterior |
| Temperatures mean | 0.500 | 0.979 | 0.500 | 0.984 | 0.500 | 0.972 | 0.500 | 0.953 |
| Policy Lags | 0.500 | 0.885 | 0.500 | 0.872 | 0.500 | 0.844 | 0.500 | 0.799 |
| Relative Humidity | 0.500 | 0.436 | 0.500 | 0.576 | 0.500 | 0.577 | 0.500 | 0.559 |
| Tests | 0.500 | 0.298 | 0.500 | 0.459 | 0.500 | 0.477 | 0.500 | 0.463 |
| Health spending | 0.500 | 0.169 | 0.500 | 0.250 | 0.500 | 0.293 | 0.500 | 0.327 |
| Social Mobility | 0.500 | 0.138 | 0.500 | 0.246 | 0.500 | 0.286 | 0.500 | 0.315 |
| Share of old population | 0.500 | 0.138 | 0.500 | 0.260 | 0.500 | 0.298 | 0.500 | 0.322 |
| Air Pollution | 0.500 | 0.119 | 0.500 | 0.214 | 0.500 | 0.250 | 0.500 | 0.277 |
| Share of young population | 0.500 | 0.118 | 0.500 | 0.216 | 0.500 | 0.256 | 0.500 | 0.284 |
| Population density | 0.500 | 0.117 | 0.500 | 0.215 | 0.500 | 0.257 | 0.500 | 0.289 |

Notes: The dependent variable in all regressions is the total number of contagions per 100,000 inhabitants from the Italian MCP. The results reported correspond to the estimation of all the models in the model space including any combination of the 10 variables.

We first check the sensitivity of our results to the the definition of the Binomial prior on the model space, with *φ* = 0.5, which implies APIPs for the variables of the 50%. Thus, we depart from the baseline specification and we set the prior model size to 6, 7 and 8 regressors respectively, by adjusting *φ* in each case. As shown in Table (4), the effect of increasing the prior model size has a stronger effect on the PIPs than the g-prior, since the employment of priors favoring large model sizes increases slightly the PIPs of most of the potential determinants.

However, when comparing the APIPs of each variable with the estimated PIPs implied by the data, we do not find any significant change in our previous results. Temperatures and policy lags remain to be the most robust determinants of contagion differentials with PIPs that are always above the 90%. The only difference appears if one holds an priori view of COVID-19 contagions being driven by a larger set of potential variables, for prior model sizes equal to 7 and 8 (i.e, assuming variables have APIPs of the 70 and 80% respectively). In this context, one should also consider the relative humidity as a relevant driver of contagion differentials. However, the result of a categorization of all the other variables as “unimportant factors” does not change.

Secondly, we consider a variety of different g-priors specifications on the parameters while holding fixed *φ* = 0.5. These prior-distributions differ from our baseline g-prior, the BRIC which sets *g* = *max N*; *K*^2^. In this group of g-prior distributions we consider the (i) Unit information prior (UIP) which sets *g* = *N*; the (ii) Risk information criteria prior (RIC) where *g* = *K*^2^ and (iii) Empirical Bayes prior (EBL) which is a model *k* specific g-prior estimated via maximum likelihood. In this case *g* = *max* (0*, F_k_*) where 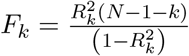. Finally, we consider the Hyper-g prior which relies on a Beta prior on the shrinkage factor of the form 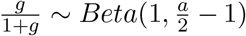 where in this specific case, *a* = 3.5.

In this context, temperatures and policy lags always appear to be the most important variables. As regards the role of relative humidity, we find that when employing UIP, EBL or Hyper-g priors relative humidity displays PIPs above the APIPs. Taken together the results of these robustness checks suggest that the main results are not driven by prior information and that only if one is willing to accept more candidate determinations as a part of the explanation of cross-regional differentials the role of humidity may change.

#### 6.2.2 Frequentist weights

We now investigate if a frequentist model averaging produces similar results. Despite the similarity in spirit and objectives between Frequentist Model Averaging (FMA) and BMA, both techniques differ in their approach to inference (Moral-Benito, 2015). As explained by Steel (2017), FMA analysis tend to focus on estimators and their properties, and do not require a prior on the parameters. In FMA, the parameters are treated as fixed, yet unknown, and are not assigned any probabilistic interpretation associated with prior knowledge or learning from data. To date, the empirical literature of FMA has mainly focused on forecasts combinations (Steel, 2017). However, as we show below, this approach can also be employed to derive metrics of great interest to analyze the importance of the potential drivers of COVID-19 differentials. Let the FMA estimator of *η* be given by:

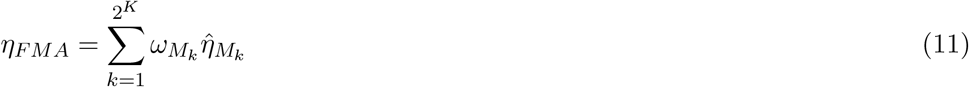

where 0 ≤ *ω_M_k* ≤ 1 and 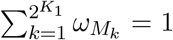 such that it integrates both, model selection and parameter estimation. Following Buckland *et al*. (1997) the estimated standard error in FMA is given by:

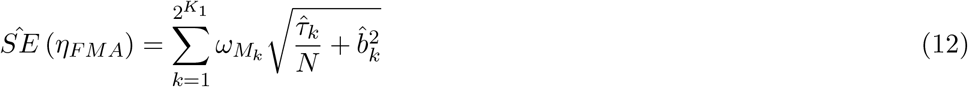

where *τ*_*k*_ is the estimated variance of the parameters in model *k* and 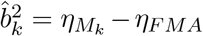 is a term that captures the uncertainty across different models. As discussed by Moral-Benito (2015) FMA estimators crucially depend on the weights selected for estimation as their asymptotic properties may differ substantially. In this analysis we consider the approach of Hansen and Racine (2012), who suggest a weighting procedure based on the minimization of a cross-validation criterion also known as the jackknife model averaging (JMA), that is appropriate for general linear models with heteroskedasticity. Cross-validated optimal weights in this context are obtained by solving:

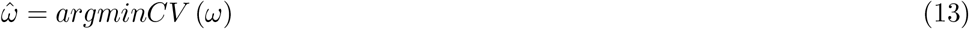

where:

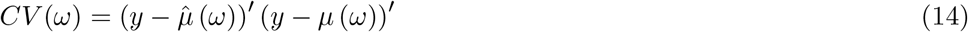

and 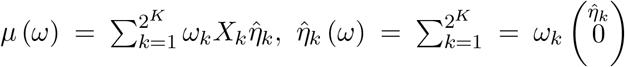 and *η* denotes the least-squared model averaging estimator of model *k*. In the FMA context it is possible to sum over model weights *ω_M_k* to derive a metric of variable importance, that is similar to that of the PIPs in the BMA, the Frequentist Variable Weight (FVW). That is, for a variable *h* we calculate the FVW as the sum of the model weights including the variable *h*:

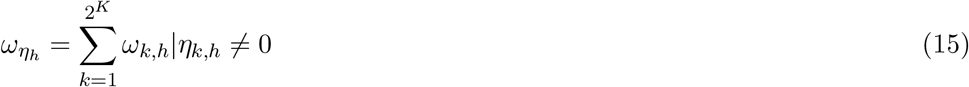

As FVWs have never been used before to asses importance, in the Online Appendix C, we provide the results of a Monte Carlo simulation study showing that variable weights *ω_η_h* can be used to generate an accurate rank on the relative importance of a regressor. In addition, to complement this metric, we also calculate the frequency of t-stats above 1.96, which corresponds to a significant covariate at the 5% level as in (Sala-i-Martin *et al*., 2004).

Table (5) shows the results derived from the jackknife model averaging analysis. Column (1) reports the FVWs, Column (2) the standardized coefficient estimate and Column (3) the fraction of models where t-stats of the variable were above 1.96, and therefore, appeared to be significant at the 5% level. The main result of temperatures being the most important determinant of COVID-19 regional contagion differentials appears is confirmed in this context as the FVW of the variable is of the 85%, clearly above the FVWs of the other predictors included. Importantly, from a frequentist inference perspective, the parameter of temperatures appears to be significant at the 5% level in all the models which reinforces our view on this variable being the most important driver. The most remarkable difference in this context is that the estimate of a 1-standard deviation shock in temperatures produces a decrease in –0.536 standard deviations of contagions, which is a slightly lower magnitude than those obtained in the BMA.

**Table 5:** Robustness checks (II): Frequentist jackknife Model Averaging.

| Variable | Jackknife<br>variable weight<br>(1) | FMA<br>Estimate<br>(2) | Fraction of<br>tstats > 1.96<br>(3) |
| --- | --- | --- | --- |
| Temperatures | 0.853 | -0.536***<br>(0.076) | 1.00 |
| Policy lags | 0.757 | 0.611***<br>(0.174) | 0.58 |
| Relative Humidity | 0.624 | -0.206*<br>(0.119) | 0.18 |
| Tests | 0.782 | -0.254*<br>(0.146) | 0.11 |
| Health spending | 0.034 | -0.029***<br>(0.008) | 0.48 |
| Share of old population | 0.147 | -0.134<br>(0.083) | 0.103 |
| Social Mobility | 0.624 | 0.079**<br>(0.035) | 0.246 |
| Share of young population | 0.051 | 0.000<br>(0.000) | 0.041 |
| Population density | 0.036 | 0.000<br>(0.000) | 0.167 |
| Air pollution | 0.403 | 0.072<br>(0.046) | 0.025 |

Notes: The results reported correspond to the estimation of all the models in the model space including any combination of the 10 variables. The jackknife (or cross-validation) choice of weight vector that solves Equation (13) is obtained using the quadprog command in MATLAB. The MATLAB codes that extend the gma function developed by Hansen and Racine (2012) to compute FVW and standard errors can be provided upon request. Standard deviations in parenthesis, \* Significant at 10% level, \*\* significant at 5% level, \*\*\* significant at 1% level.

As refers to the other variables, the results of FVWs > 0.5 in Column (1) hold for the policy lag variable, or the relative humidity. Nevertheless, in the FMA, two additional variables display FVWs > 0.5: the tests performed and the degree of social mobility of the population. The effect of testing appears to be weakly significant and negatively related to the size of the epidemic whereas that of social mobility is positive and significant at conventional 5% levels. Nevertheless, when looking at the fraction of t-stats in which these factors are significant, we find that they are only significant in the 11% and the 24.6% of the models. Therefore, models where these two variables did not appear to be relevant, were precisely those models with very large variances and noisy estimates of the parameter estimates. The opposite occurs for the health spending variable, which only obtains a FVW of the 3.4%, but its negative impact on contagions appears to be highly significant.

The results stemming from the FMA analysis confirm that temperatures are the key factor explaining differentials but interestingly, they also point out that policy variables like health-spending and social mobility, which were discarded by the PIPs in Table (2), may in indeed have played a more relevant role that previously suggested. Therefore, the results of the FMA do not alter our main result but place more emphasis in the epidemic management and actions of the policy-makers than the BMA.

#### 6.2.3 Measurement errors and definition of variables

A third concern with the validity of previous results is that measurement errors are likely to be present in our key dependent variable given that a large share of cases may have not been detected. To address this issue, we estimate the percentage of symptomatic COVID-19 cases reported in the Italian regions using case fatality ratio estimates, correcting for delays between confirmation-and-death following Russell *et al*. (2020).^16^

These authors propose to proceed in this way because of in real-time, the division of cumulative deaths (*D_t_*) by cases (*C_t_*) leads to a biased estimate of the case fatality ratio (CFR), because of this calculation does not account for (i) delays from confirmation of a case to death, and (ii) under-reporting of cases. The approach used here to obtain an accurate estimate of the size of regional infections consists in two steps. First, we use an estimated distribution of the delay from hospitalisation-to-death for known cases that are fatal, which allows us to adjust the naive estimates of the CFR to account for these delays.^17^

However, this corrected CFR (cCFR) does not account for under-reporting. Given that the best available estimates of CFR (after controlling for under-reporting) from large studies in China and South Korea are in the 1% – 1.5% range, we assume a baseline CFR of 1.4% for our analysis. Thus, if a region has a cCFR that is higher to this threshold value, it means that only a fraction of cases are being reported. In such case, the excess of the observed cCFR can be attributed to under-reporting. Specifically, under-reporting *u* of known cases is calculated for each region *i* and date *t* as:

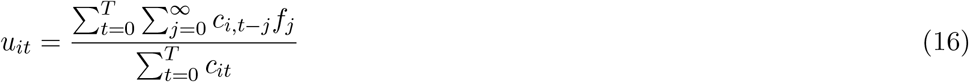

where *u_it_* represents the underestimation of the proportion of cases with known outcomes, *c_it_* is the daily incidence at time *t* and *f_t_* is the proportion of cases with delay of *t* periods between confirmation and death. We obtain the cCFR as 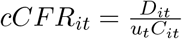 and once we have the *cCFR_it_* in hand, we estimate reporting as 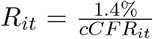. This allows us to calculate a new daily and cumulative incidence time series as:

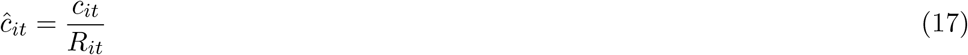

The results obtained after performing the BMA with this corrected time-series of contagions *c*_*it*_ for each region are shown in Table (6). As shown, the results using this new dependent variable confirm the temperatures are the most important determinant. In fact, when using this correction, the temperatures appear to be the only variable with a posterior probability that is above the APIP (i.e, PIP = 85.4% < APIP = 50%). The estimated posterior mean impact of an increase in one Celsius degree in this case is 21.8 times higher than that obtained when using the official data in Table (2). This can be explained by the fact that the estimated under-reporting of cases in Italian regions is quite high, ranging between the 81.45% in Lombardy and the 98.26% on Molise. Nevertheless, the standardized posterior mean estimate is of –0.613 standard deviations (in line with the baseline model) and the posterior sign positivity is 0%, which implies the statistically significant negative link is robust to all model specifications also in this context.^18^

**Table 6:**
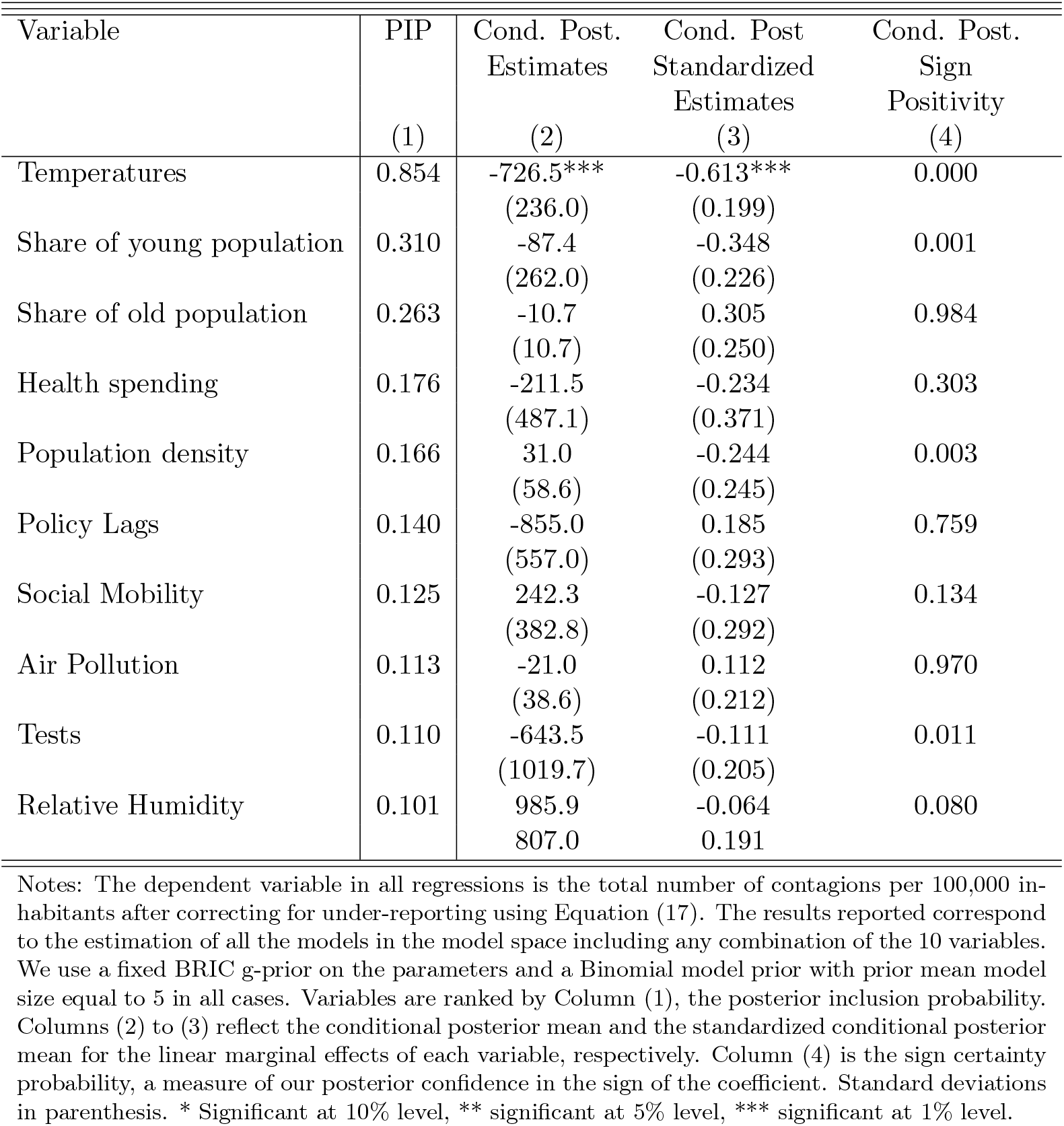
Robustness check (III): Contagions (corrected for under-reporting)

| Variable | PIP | Cond. Post.<br>Estimates | Cond. Post<br>Standardized<br>Estimates | Cond. Post.<br>Sign<br>Positivity |
| --- | --- | --- | --- | --- |
|  | (1) | (2) | (3) | (4) |
| Temperatures | 0.854 | -726.5***<br>(236.0) | -0.613***<br>(0.199) | 0.000 |
| Share of young population | 0.310 | -87.4<br>(262.0) | -0.348<br>(0.226) | 0.001 |
| Share of old population | 0.263 | -10.7<br>(10.7) | 0.305<br>(0.250) | 0.984 |
| Health spending | 0.176 | -211.5<br>(487.1) | -0.234<br>(0.371) | 0.303 |
| Population density | 0.166 | 31.0<br>(58.6) | -0.244<br>(0.245) | 0.003 |
| Policy Lags | 0.140 | -855.0<br>(557.0) | 0.185<br>(0.293) | 0.759 |
| Social Mobility | 0.125 | 242.3<br>(382.8) | -0.127<br>(0.292) | 0.134 |
| Air Pollution | 0.113 | -21.0<br>(38.6) | 0.112<br>(0.212) | 0.970 |
| Tests | 0.110 | -643.5<br>(1019.7) | -0.111<br>(0.205) | 0.011 |
| Relative Humidity | 0.101 | 985.9<br>807.0 | -0.064<br>0.191 | 0.080 |

Notes: The dependent variable in all regressions is the total number of contagions per 100,000 inhabitants after correcting for under-reporting using Equation (17). The results reported correspond to the estimation of all the models in the model space including any combination of the 10 variables. We use a fixed BRIC g-prior on the parameters and a Binomial model prior with prior mean model size equal to 5 in all cases. Variables are ranked by Column (1), the posterior inclusion probability. Columns (2) to (3) reflect the conditional posterior mean and the standardized conditional posterior mean for the linear marginal effects of each variable, respectively. Column (4) is the sign certainty probability, a measure of our posterior confidence in the sign of the coefficient. Standard deviations in parenthesis. \* Significant at 10% level, \*\* significant at 5% level, \*\*\* significant at 1% level.

Another issue that deserves attention and that might affect the validity and interpretation of our previous results is that of the measurement of temperatures, as it could be that the maximum or the minimum daily temperature are those favoring or hampering viral transmission. Although they are correlated with the mean temperatures they imply upper and lower bounds, producing a different impact on contagions.

Table (7) reports the results obtained when using these two measurements of temperatures. We use the average maximum and minimum temperatures 2 meters above the surface using the centroid of the region using data taken from the NASA-POWER v8 GIS database. The results of the benchmark analysis are shown in Columns (1) to (3) for the sake of comparison, while Columns (4) to (6) and Columns (7) to (9) display those obtained for the minimum and maximum temperatures respectively. As observed, both minimum and maximum temperatures also appear to be the more relevant than the other determinants, with PIPs of 95.2% and 99.4%. In either case, the estimated standardized coefficient is always negative across all model specifications in the range of –0.66 to –0.71 standard deviations. This means that the most extreme daily values do not alter our results.

**Table 7:**
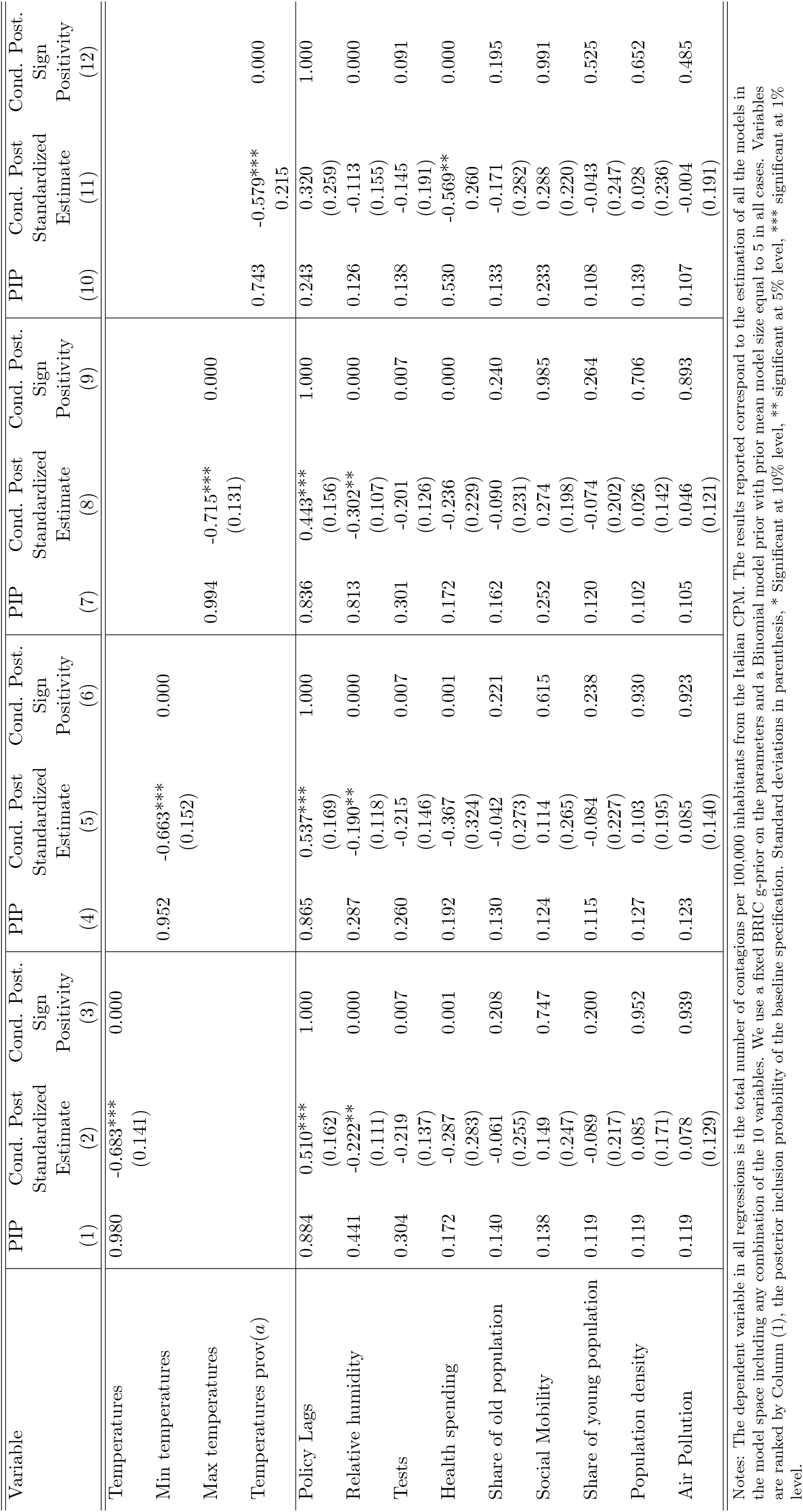
Robustness checks (IV): Alternative measurements of temperatures.

To complement this information we use a different metric of temperatures, obtained from the online climate portal “ilmeteo.it”, which reports historical monthly measurements of minimum and maximum temperatures for different provinces and meteorological stations scattered within the boundaries of each of the regions. This metric departs from the meteorological measurement performed at the centroid of the region given that in this case, the average maximum and minimum temperatures for all the spatial data points within each region are used to produce a regional estimate of mean temperatures. Again, when using this alternative measurement of temperatures, we find it to be the most relevant one, with a PIP of the 74.3%. Therefore, the main results reported in Table (2) are robust to these alternative measurement.

#### 6.2.4 Additional regressors

As mentioned above, our baseline specification includes different controls based on the literature analyzing COVID-19 differentials. However, one may still argue that the effect of temperatures on the dependent variable could simply be capturing the effect of some omitted correlated determinant of contagions. In order to investigate further this issue, we now extend our baseline specification by including different covariates that may be associated with both, temperatures and contagions at the regional level.

In particular, we begin by considering the role played by the distance to the epicenter of the epidemic, which in Italy was located in Lombardy. In an analogy with the diffusion of the impacts of an earthquake, we expect that a higher regional distance to the epicenter of the pandemic may have reduced the share of imported contagions and as a consequence, the final epidemic size. This intuition is supported by the work of Fang and Wahba (2020), where distance to Wuhan is observed to be an important predictor of contagion differentials among Chinese cities. In addition, controlling for distances to Lombardy is relevant in the Italian context, given that regions in the south of the country or in the Mediterranean islands far away from Lombardy, tend to have better climatic conditions.

We also control for the presence of comorbidities in the population as there is evidence that suggests that COVID-19 infections exerts a higher impact on those suffering from other health problems, such that their clinical evolution is usually worse (Guan *et al*., 2020). To account for these differentials, we aggregate into a composite index the annual number of patients discharged by hospitals with either respiratory diseases, neoplasms and diseases of the circulatory system.

In addition, we consider differences in the regional the quality of government. Previous studies have shown that regional quality of government differentials are a robust determinant explaining regional resilience during the downturn of the Great Recession (Rios and Gianmoena, 2020). In the context of a health crisis, a higher quality of government is expected to influence not only the provision and the quality of health-care services but also the adaptability of institutions to a changing and turbulent environment, which in turn may have helped to buffer the epidemic shock. Hence, to control for regional differentials across regions in the quality of government, we use the composite indicator developed by (Charron *et al*., 2014).^19^

Moreover, as discussed in Section (2), it is possible that what matters for SARS-CoV2 viral transmission are not the temperatures per se, but the levels of sunlight and UV radiation, which are deemed to degrade viral genetic material and increase host defenses through enhanced production of vitamins (Canell *et al*., 2006; Martineau *et al*., 2017). Because of there is a strong correlation between regional temperatures and solar radiation, we disentangle the effects of these two variables by including the mean solar radiation flux in the centroid of the region by using data taken from the NASA-POWER-v8 database.

Finally, we consider the potential effect of infrastructure density, measured as the number of road and railway kilometers per squared kilometer. This variable is expected to capture differences in the degree of social mobility and connectedness within the region. The reason for including this variable is that despite the fact that the first outbreak could have struck anywhere, regions with large cities like Milan or Rome should have a bias to attracting them, mainly because they have commercial hubs and large transport nodes with a massive influx of tourism and travelers. Likewise, the use of public transport may have been a key factor in the rapid spread of COVID-19 (AQR, 2020b).

Table (8) show the results obtained when the BMA analysis is performed again including these additional controls. As can be seen, only the temperatures, the policy lags, the distance to Lombardy and the relative humidity display statistically significant effects at the 5% level. Moreover, we find that the most relevant factor are the regional temperatures with a PIP of the 92.9% and with a coefficient that continues to be negative and statistically significant in all cases. Therefore, the newly-added controls do not alter the main result of the paper.^20^

**Table 8:** Robustness checks (V): Extended set of regressors.

| Variable | PIPs | Cond. Post.<br>Standardized<br>Estimate | Cond.Post.<br>Sign<br>Positivity |
| --- | --- | --- | --- |
|  | (1) | (2) | (3) |
| Temperatures | 0.929 | -0.587***<br>(0.182) | 0.000 |
| Policy Lags | 0.538 | 0.471***<br>(0.190) | 0.994 |
| Distance to Epicenter | 0.478 | -0.548**<br>(0.235) | 0.000 |
| Relative Humidity | 0.311 | -0.208**<br>(0.113) | 0.000 |
| Tests | 0.180 | -0.190<br>(0.145) | 0.007 |
| Health spending | 0.117 | -0.226<br>(0.332) | 0.254 |
| Social Mobility | 0.098 | 0.112<br>(0.240) | 0.754 |
| Share of old population | 0.097 | -0.040<br>(0.235) | 0.392 |
| Air Pollution | 0.091 | 0.021<br>(0.163) | 0.577 |
| Population density | 0.090 | 0.054<br>(0.179) | 0.873 |
| Share of young population | 0.088 | -0.039<br>(0.222) | 0.334 |
| Quality of government | 0.082 | 0.019<br>(0.249) | 0.389 |
| Comorbidities | 0.080 | -0.029<br>(0.169) | 0.279 |
| Solar radiation | 0.079 | -0.039<br>(0.228) | 0.446 |
| Infrastructure density | 0.078 | 0.019<br>(0.158) | 0.599 |

Notes: The dependent variable in regressions is the total number of contagions per 100,000 inhabitants. The results reported correspond to the estimation of the top 100 models from all the possible regressions including any combination of the 15 variables. We use the reversible jump $MC^3$ sampler algorithm implemented by Zeugner and Feldkircher (2015) in R. We use a fixed BRIC g-prior on the parameters and a Binomial model prior with prior mean model size equal to 7.5. Variables are ranked by Column (1), the posterior inclusion probability. Standard errors in parenthesis, \* Significant at 10% level, \*\* significant at 5% level, \*\*\* significant at 1% level.

#### 6.3 Alternative Variable Importance Metrics

The results obtained so far imply that temperatures are the most robust determinant of cross-regional contagion differentials in Italy. We now investigate this issue from a different perspective. As a complement to the Bayesian and the Frequentist Model Averaging approaches developed before, we explore the relative importance of the various factors that could affect contagions by means of methods that partition the proportion of explained model’s variance among multiple predictors (i.e, the *R*^2^ of the model). The term relative importance here refers to the contribution a variable makes to the prediction of contagions by itself and in combination with other predictor variables, which differs from classical statistical inference where a variable may explain only a small proportion of predictable variance and yet be considered very meaningful (Johnson and LeBreton, 2004). To understand the intuition behind this approach recall that in the context of a cross-sectional regression, with *n* = 1*, …, N*, the *R*^2^ informs on the model’s explained variability across spatial units (regions). Thus, decompositions on the relative importance of a factor *X^k^*, tell us the percentage of explained disparities across spatial units due to factor *k*.

In a linear regression framework with *K* explanatory factors the variance of the model is given by:

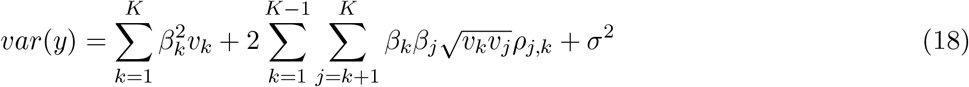

where *β_k_* stands for the parameter of variable *k, v_k_* for the regressor variance, *ρ_j,k_* is the inter-regressor correlations and *σ*^2^ denotes the unexplained variance. As explained by Gromping (2007) variable importance methods that decompose *R*^2^, have to decompose the explained variance of the model (i.e, the first two summands above). However, these variances can only be uniquely partitioned in the case of uncorrelated regressors. Whenever *ρ_j,k_* ≠ 0, different methods lead to different results.

Given that the increase in the *R*^2^ allocated to a certain factor *X_k_* depends on which other variables are already in the model when *X_k_* is added to be part of the explanation, Gromping (2007) propose to use the Lindeman-Merenda-Gold method which consists on averaging such order-dependent *R*^2^ allocations over all *p*! orderings to produce a fair unique assessment of importance. The Proportional Marginal Variance Decomosition (PMVD) approach proposed by Feldman (2005), is a modified version with data dependent weights penalizing variables that are not statistically significant. Others, like Zuber and Strimmer (2010) and Zuber and Strimmer (2011) have proposed to by-pass the need of estimating all possible models by decorrelating the variables with a Mahalanobis transformation and suggest to look directly at Correlation-Adjusted (marginal) coRelation (CAR) scores, whereas Genizi (1993) proposed a similar metric that can be viewed as the weighted average of squared CAR scores.

To study the relative contribution of the various factors affecting contagion, we employ the LMG method (Gromping, 2007), the PMVD (Feldman, 2005) and the Genizi and the CAR scores (Genizi, 1993; Zuber and Strimmer, 2010 Zuber and Strimmer, 2011). The formulas employed to obtained the decompositions implied by these metrics are detailed in the Appendix D. Table (9) reports the estimated relative importance metrics for each group of variables and each variable alone.

**Table 9:**
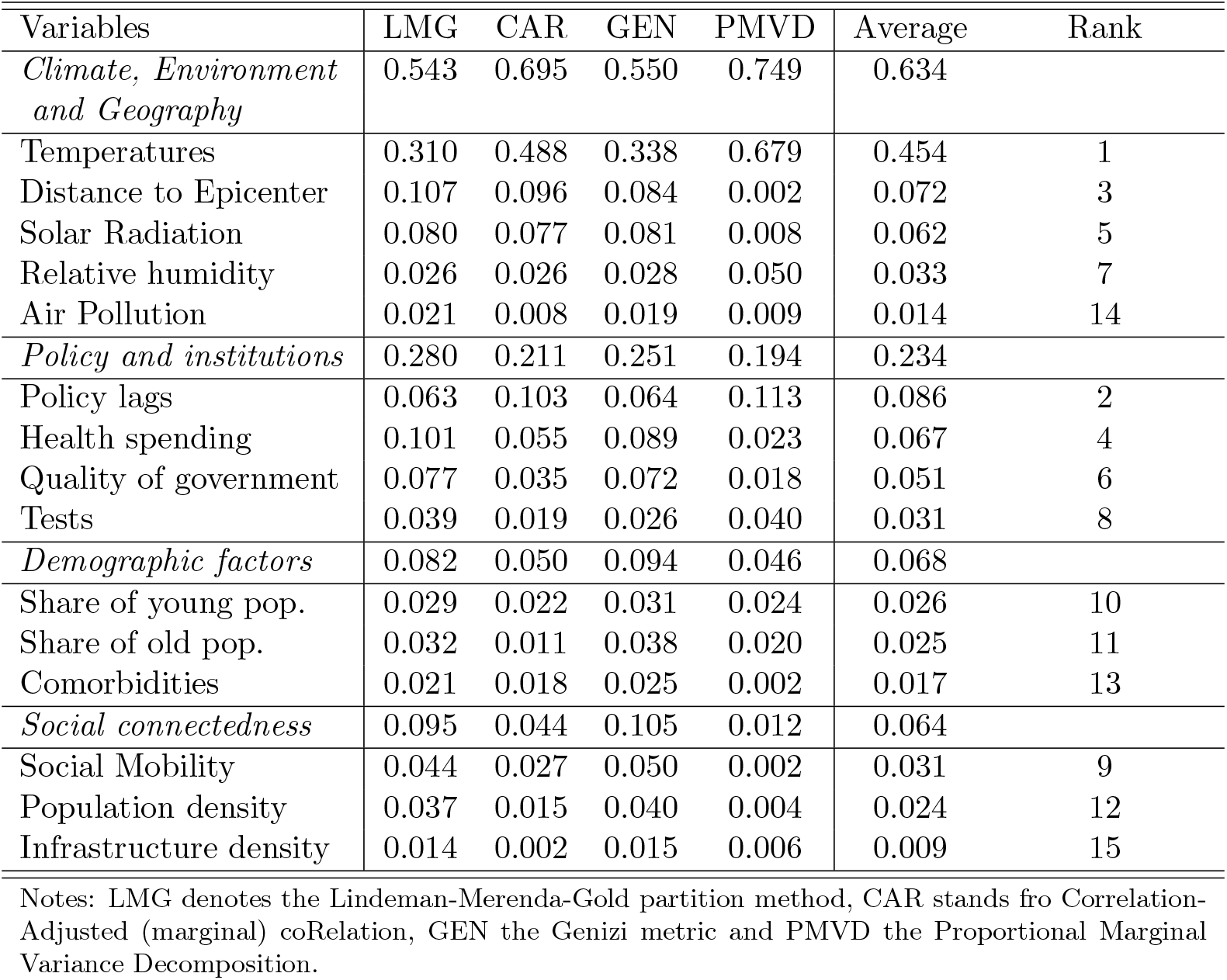
Regression Relative Importance Metrics.

As can be seen, the variables that belong to the climate, environmental and geography factors explain 63.4% of the variability in contagions. Within this group, leaving the role of temperatures aside, the distance to the epicenter of the epidemic outbreak (7.2%) and solar radiation (6.2%) are the most important ones. Relative humidity differentials just explain a 3.3% of the variability in contagions which contrasts with the higher importance attributed to this variable in the BMA and FMA approaches. However, the low importance attributed by the relative importance analysis to the variability in air pollution (1.4%) is in agreement with previous approaches.

The second group of factors in terms of importance are the institutional factors and those related to the policy-response to the epidemic which all together, explain a 23.4% of disparities. The individual rankings of all the factors included in these group are above demographic and social connectedness ones. After the policy lag variable which is ranked in the second place, the share of health spending in the GDP (6.7%) is ranked as the fourth variable wheres the quality of regional government (5.1%) in the sixth place, explain a non-negligible share of contagion variation. The tests performed relative to the population are ranked in the eighth place, explaining the 3.31% of the model’s fit to the data.

In a much lesser degree of importance we find the demographic characteristics (6.8%) and the social connectedness factors (6.4%). Among the demographic variables, the shares are quite equi-distributed since the share of young population (2.6%), the share of old population (2.5%) and the regional comorbidity index (1.7%) a similar portion of the *R*^2^. Regarding the factors that capture regional differences in social connectedness the most prominent variable is the degree of social mobility (3.1%) above the population density (2.4%) and the infrastructure density (0.9%).

Taken together, the results suggest that although institutional settings and political interventions have been important, regional contagion outcomes in Italy have been largely determined by factors outside the scope of regional policy-makers.

### 7 Conclusions and Policy Implications

This study has examined the determinants of regional COVID-19 contagion differentials in Italy during the period ranging from February 24 to April 15 covering the first wave of the epidemic outbreak. They key contribution of this analysis is methodological given that we consider the effect of a greater number of determinants than previous studies and we employ BMA techniques to account for model uncertainty in a cross-regional contagion regression framework.

Within the BMA, we compute the PIPs for the different indicators to generate a probabilistic ranking of relevance for the various contagion determinants. The analysis reveals the temperatures are the top determinant shaping regional reactions to the epidemic outbreak in Italy with a PIP of the 97.9%. We find that the negative and statistically significant effect on viral transmission is robust to all the model specifications. The most plausible explanation for this link is that lower temperatures may have increased the efficiency of viral propagation decreasing host defenses and favoring the conditions for aerosol transmission in closed spaces. The result of a negative relationship between temperatures and contagions is robust to (i) different forms of prior distributions elicitation, (ii) the procedure to assign importance-weights to the regressors, (iii) the presence of measurement errors in official data due to under-reporting, (iv) the employment of alternative definitions of temperatures, or (v) the inclusion of additional covariates.

In a second step, we calculate relative importance metrics that allow to perform an accurate partitioning of the model’s fit to the data. The results show that climate and environmental factors explain 63% of regional differences. Specifically, we find that temperatures alone are responsible for the 45.4% of the observed disparities in contagions. The set of factors related to the institutional setting and the policy response to the epidemic crisis appear in a second level of importance, whereas factors related to the degree of social connectedness or the demographic characteristics are less relevant in this context.

Although the limited spatial coverage of the analysis implies that our main findings should be taken with caution and cannot be extrapolated elsewhere, the results of the paper raise potentially important policy implications, especially at a time in which there is an active public debate on the most appropriate instruments to reduce the impact of the epidemic outbreak on regional economies.

First, although our analysis suggests that higher temperatures may have contributed to decrease the viral transmission efficiency, this does not imply that contagions will not occur during the summer season or in warmer environments. In fact, given the wide range of average temperatures in which contagions occurred in Italy, from –4 to 10 Celsius degrees, our findings suggest that transmission is likely to occur in diverse environments. The strong importance that our modeling exercise attributes to temperatures and delays in lockdown policies, indicates that in the absence of a vaccine, the combined effect of a relaxation of the lockdown measures and the expected drop in temperatures during the fall and/or the following winter, could re-create the conditions for a new epidemic outbreak.

It is precisely, in the hypothetical context of second epidemic wave, where the results obtained here regarding the role played by some policy related variables may be useful and help to guide policy-making.

An interesting result, revealed by the posterior jointness analysis, is that there is an important degree of substitutability between temperatures, lockdowns, and the regional health-spending on GDP. Therefore, the negative effect on contagions exerted by health-spending and the fact that in the absence of lockdowns this variable would have been considered as much more relevant from a probabilistic point of view, can be interpreted as evidence in favor of the need to increase the share of economic resources devoted to robustify the health-care system. Since regional lockdowns are very costly from an economic point of view, our results point to health-care system investments as a less costly but effective policy to deal with the epidemic.

A similar implication can be derived for the degree of social mobility as it has been observed it tends to increase contagions. Given that this variable also appears to be substitute of lockdowns, policies aiming at reducing the level of social mobility, as it is the case of adopting teleworking (working from home), could help to reduce daily commutes and the significant possibility of transmission in office environments. Thus, the expansion of teleworking may help reduce costs to the economy and society.

Finally, we would like to highlight some future empirical avenues of research that have a strong potential to gain a deeper understanding on the linkages between climate and COVID-19 contagions. From the methodological point of view, one option is the extension of the BMA framework employed here to the context of a dynamic spatial panel data model including fixed effects (Lee and Yu, 2010) or adaptive fixed effects (Shi and Lee, 2017). This could allow to investigate probabilistic importance by exploiting not only cross-regional variation but also time-variation while controlling for unobserved heterogeneity. A second avenue of research consists in the development of a homogeneous and comparable cross-country regional dataset of contagions. This type of study will require the employment of correction methods such as the one we have implemented here. In the European context, high-frequency mortality monitoring data could allow to correct under-reporting in the deaths, which could be useful to further refine the methodology of Russell *et al*. (2020).

## Data Availability

The data, MATLAB and R codes can be provided upon request.

## 8 Appendix

## Appendix A: Additional Figures and Tables

**Figure A1:**
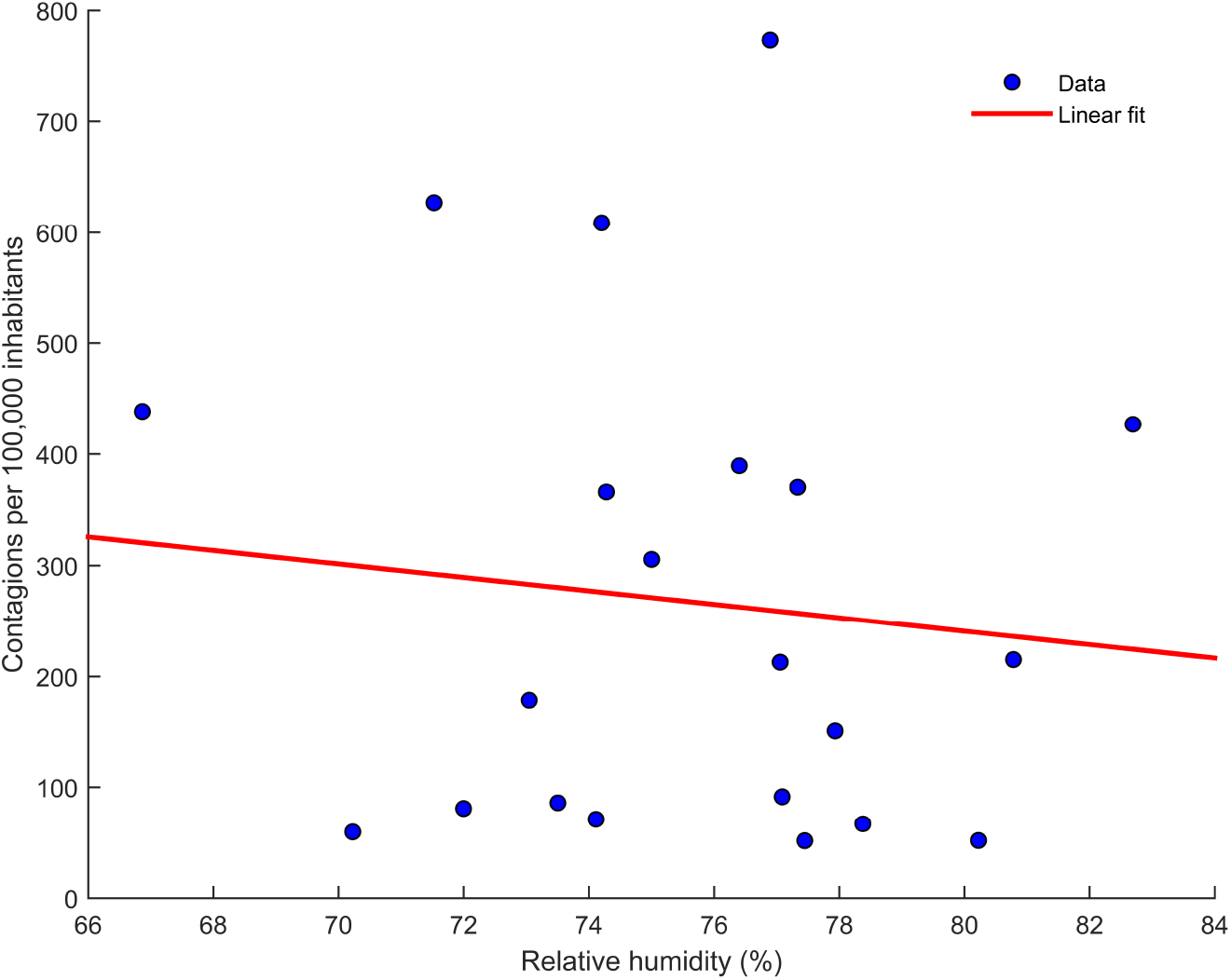
The Link between Relative Humidity and Contagions

**Figure A2:**
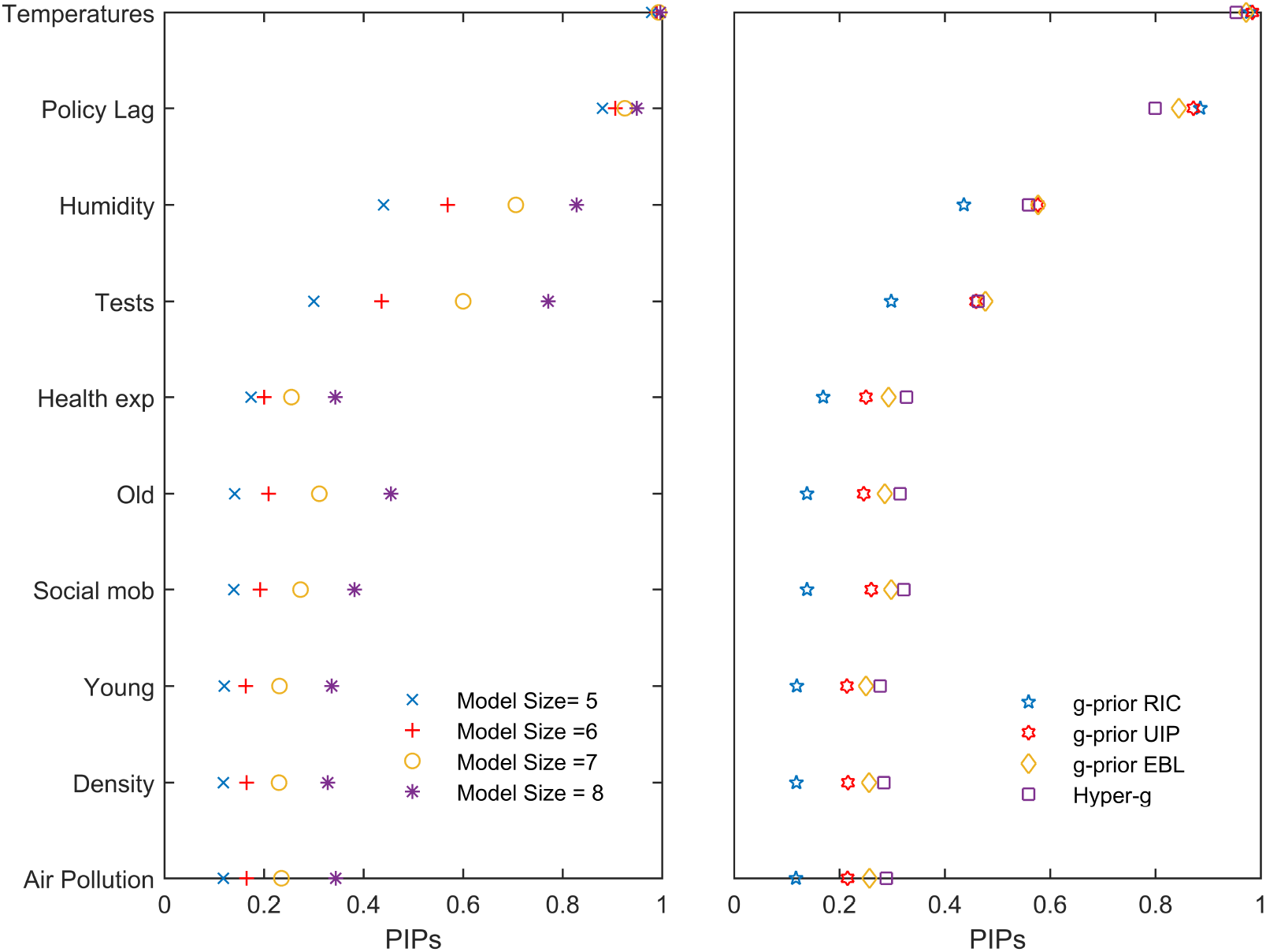
The role of g-priors and model size priors

**Figure A3:**
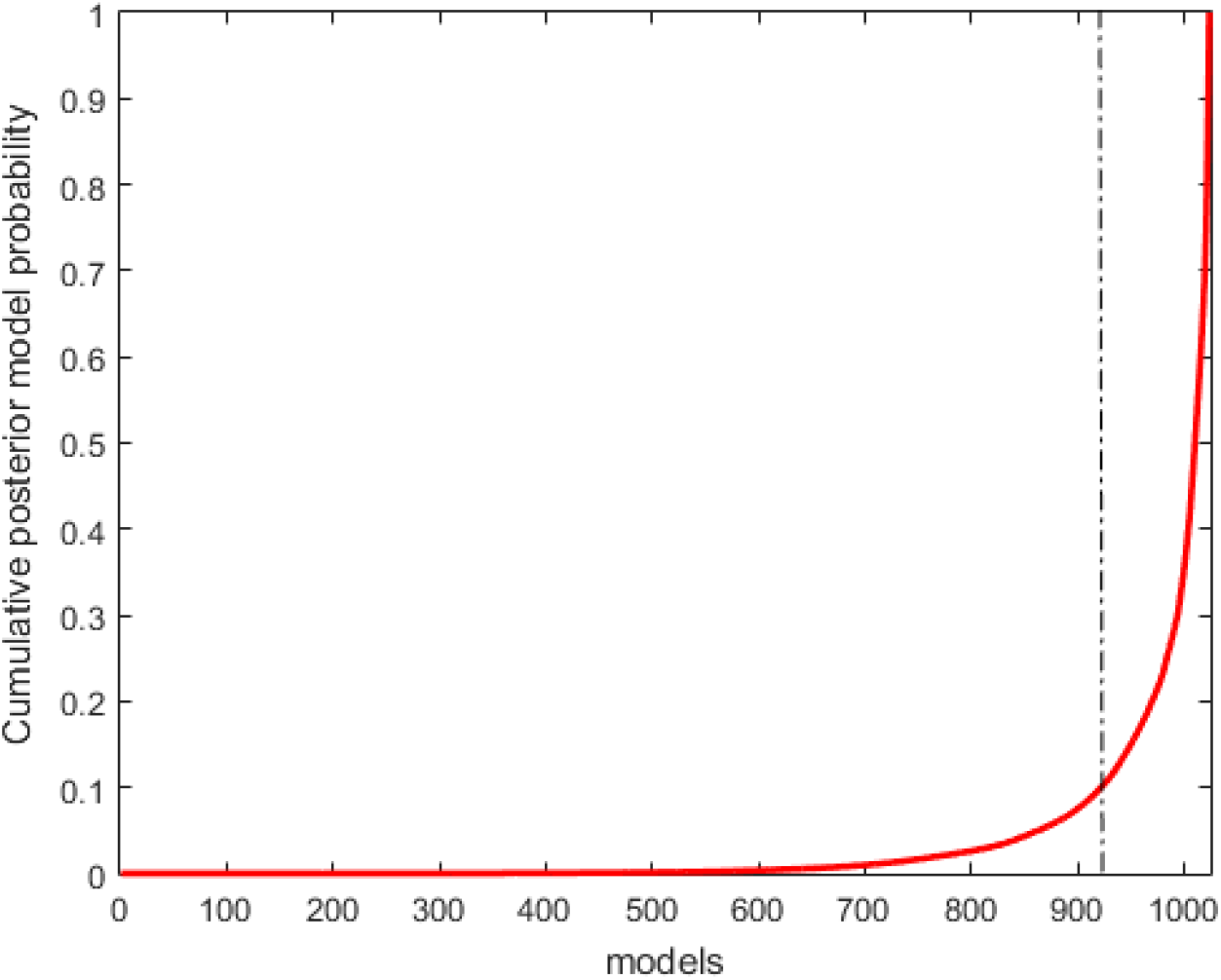
Cumulative Posterior Model Probabilities

**Table A1:** Corrected Contagion Estimates vs Official Data

| Code | Region | Official<br>New cases<br>at the peak | Estimated<br>New cases<br>at the peak | Official<br>Final epidemic<br>size | Estimated<br>Final epidemic<br>size | Estimated<br>Cumulative<br>under-reporting |
| --- | --- | --- | --- | --- | --- | --- |
| ITC1 | Piemonte | 996 | 18,423 | 19108 | 289,885 | 93.41% |
| ITC2 | Valle d'Aosta | 73 | 3,556 | 971 | 29,388 | 96.70% |
| ITC3 | Liguria | 262 | 12,500 | 6039 | 132,476 | 95.44% |
| ITC4 | Lombardia | 3251 | 14,315 | 63094 | 340,135 | 81.45% |
| ITF1 | Abruzzo | 160 | 2,536 | 2346 | 33,094 | 92.91% |
| ITF2 | Molise | 30 | 3,000 | 263 | 15,097 | 98.26% |
| ITF3 | Campania | 225 | 3,056 | 3887 | 41,970 | 90.74% |
| ITF4 | Puglia | 163 | 6,844 | 3258 | 47,883 | 93.20% |
| ITF5 | Basilicata | 31 | 1,153 | 336 | 12,877 | 97.39% |
| ITF6 | Calabria | 101 | 2,897 | 1009 | 29,540 | 96.58% |
| ITG1 | Sicilia | 170 | 6,010 | 2579 | 43,310 | 94.05% |
| ITG2 | Sardegna | 94 | 10,494 | 1164 | 33,860 | 96.56% |
| ITH1 | Provincia Autonoma di Bolzano | 141 | 2,275 | 2267 | 35,200 | 93.56% |
| ITH2 | Provincia Autonoma di Trento | 172 | 5,677 | 3294 | 42,439 | 92.24% |
| ITH3 | Veneto | 586 | 12,508 | 14990 | 221,530 | 93.23% |
| ITH4 | Friuli-Venezia Giulia | 147 | 2,837 | 2616 | 45,656 | 94.27% |
| ITH5 | Emilia-Romagna | 980 | 4,598 | 21486 | 147,591 | 85.44% |
| ITI1 | Toscana | 406 | 20,177 | 7943 | 152,756 | 94.80% |
| ITI2 | Umbria | 92 | 1,673 | 1329 | 17,977 | 92.61% |
| ITI3 | Marche | 268 | 15,450 | 5582 | 167,750 | 96.67% |
| ITI4 | Lazio | 210 | 7,129 | 5380 | 62,385 | 91.38% |

Notes: Official estimates are taken from the Ministry of Civil Protection, whereas corrected estimates are obtained using the methodology outlined by Russell *et al.* (2020).

**Table A2:** Data, variable definitions and sources

| Variable | Variable definition | Source | Date |
| --- | --- | --- | --- |
| Contagions | Total COVID-19 contagions per 100,000 inhabitants in April 15 | MCP | 2020 |
| Contagions (corrected) | Total COVID-19 contagions per 100,000 inhabitants in April 15 corrected for under-reporting as in Russell *et al.* (2020) | Own elaboration | 2020 |
| **A. Climate, Environment and Geography** |  |  |  |
| Mean temperatures(a) | Average mean daily temperature at 2 meters above the surface during February 2020 | NASA-POWER v8 | 2020 |
| Minimum temperatures(a) | Average minimum daily temperature at 2 meters above the surface during February 2020 | NASA-POWER v8 | 2020 |
| Maximum temperatures(a) | Average maximum daily temperature at 2 meters above the surface during February 2020 | NASA-POWER v8 | 2020 |
| Mean temperatures - prov | Average mean daily temperature during February 2020 across provincial meteorological stations | ilmeteo | 2020 |
| Relative humidity(a) | Average of the mean daily relative humidity at 2 meters above the surface during February 2020 | NASA-POWER v8 | 2020 |
| UV Radiation(a) | Average of the mean daily solar radiation incident on a horizontal surface for all sky conditions | NASA-POWER v8 | 2020 |
| Air pollution | Percentage of population exposed to more than 15 micrograms/m<sup>3</sup> of PM<sub>2.5</sub> particles | OECD | 2020 |
| Distance to the epicenter | Distance in kilometers from Lombardy (regional centroid distances) |  |  |
| **B. Economic and Political factors** |  |  |  |
| Policy Lags | Number of days elapsed between 1 infections per 100,000 inhabitants and the lockdown | Own elaboration | 2020 |
| Health expenditure | Health spending (in % of GDP) | Statista, Eurostat | 2019 |
| Quality of government | Regional Quality of government index based on the indicators of corruption, regulatory quality, and impartiality | Charron *et al.* (2014) | 2017 |
| Tests | Number of COVID-19 tests performed per 100,000 inhabitants | MCP | 2020 |
| **C. Demographic factors** |  |  |  |
| Share of young | Share of population aged between 0 and 35 years old | Eurostat | 2019 |
| Share of very old | Share of population aged above 75 years old | Eurostat | 2019 |
| Comorbidities | Hospital discharges of patients diagnosed with (i) respiratory diseases, (ii) Neoplasms and (iii) Diseases of the circulatory system (per 100,000 inhabitants) | Eurostat | 2018 |
| **D. Social connectedness** |  |  |  |
| Infrastructure density | Number of kilometres of motorways and railways network per squared kilometer | Eurostat | 2018 |
| Population density | Population per squared kilometer | Eurostat | 2018 |
| Social Mobility Index | Average municipal commuting flows (incoming and outgoing) relative to the population employed | Pluchino et al. (2020) | 2020 |

Notes: MCP Ministry of Civil Protection, NASA-Prediction Of Worldwide Energy Resources (NASA-POWER) v8 GIS database.

**Table A3:**
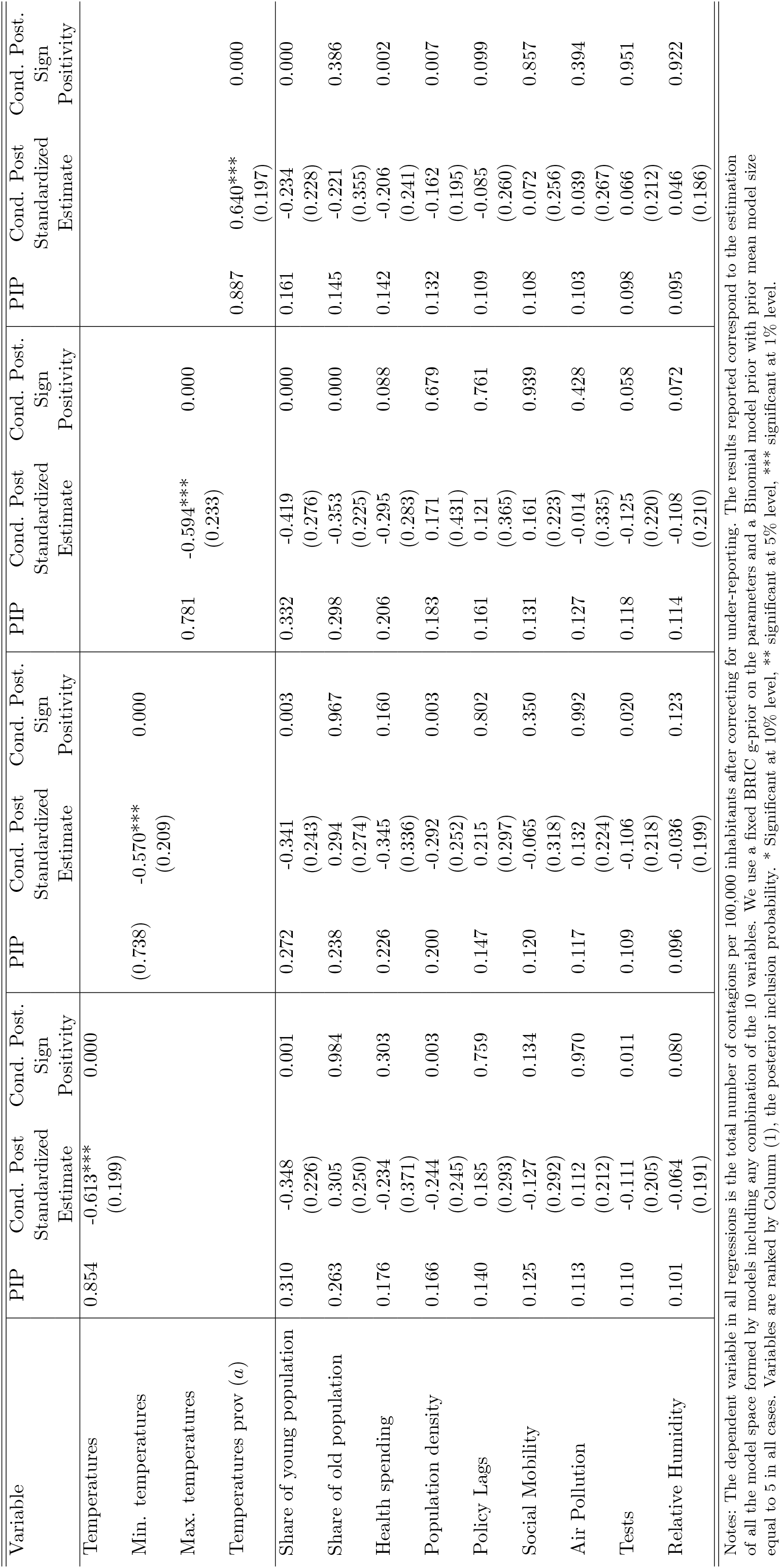
Corrected contagions and alternative measurements of temperatures

**Table A4:**
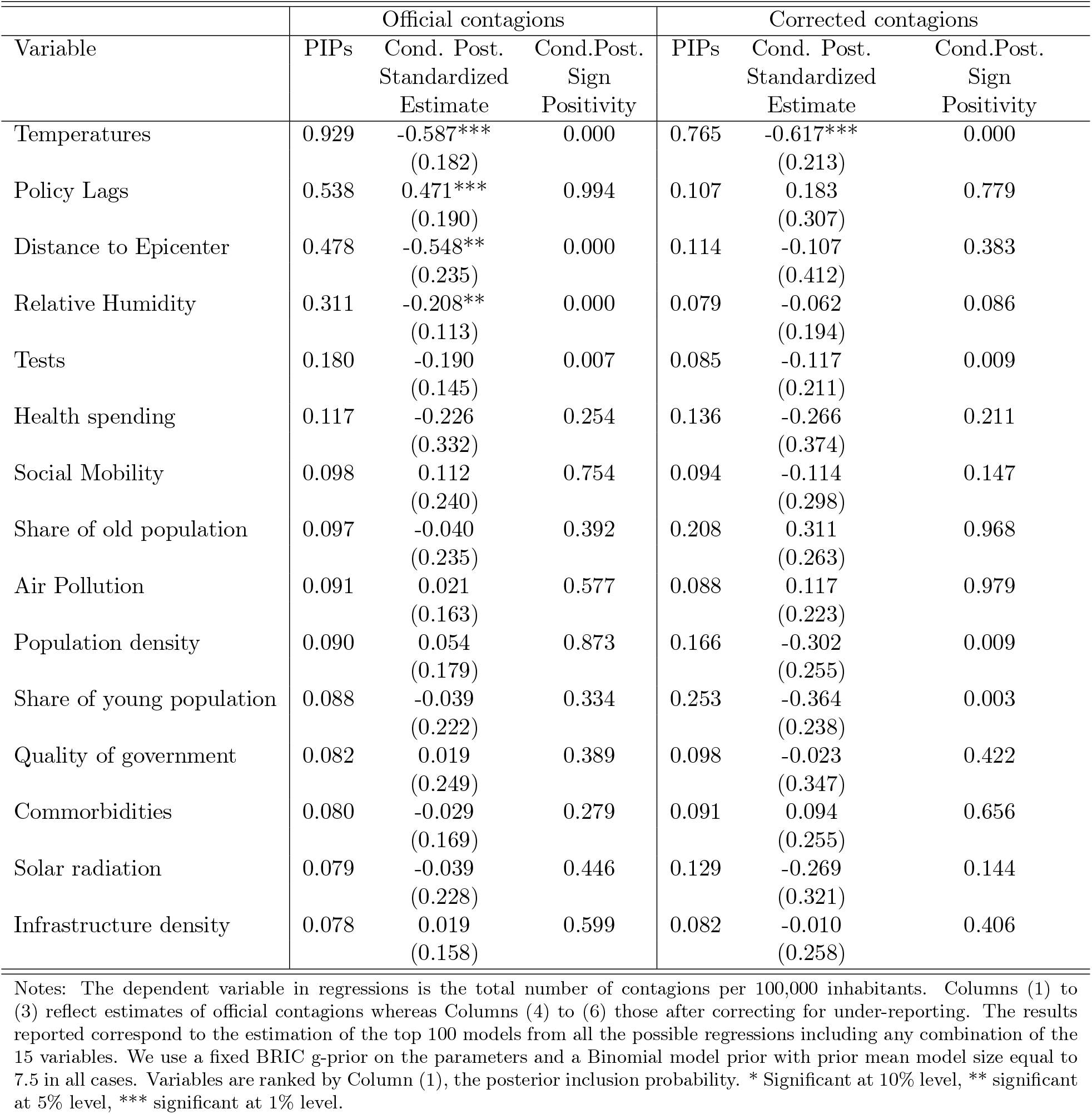
Extended set of regressors, Official vs Corrected

| Variable | Official contagions |  |  | Corrected contagions |  |  |
| --- | --- | --- | --- | --- | --- | --- |
|  | PIPs | Cond. Post.<br>Standardized<br>Estimate | Cond.Post.<br>Sign<br>Positivity | PIPs | Cond. Post.<br>Standardized<br>Estimate | Cond.Post.<br>Sign<br>Positivity |
| Temperatures | 0.929 | -0.587***<br>(0.182) | 0.000 | 0.765 | -0.617***<br>(0.213) | 0.000 |
| Policy Lags | 0.538 | 0.471***<br>(0.190) | 0.994 | 0.107 | 0.183<br>(0.307) | 0.779 |
| Distance to Epicenter | 0.478 | -0.548**<br>(0.235) | 0.000 | 0.114 | -0.107<br>(0.412) | 0.383 |
| Relative Humidity | 0.311 | -0.208**<br>(0.113) | 0.000 | 0.079 | -0.062<br>(0.194) | 0.086 |
| Tests | 0.180 | -0.190<br>(0.145) | 0.007 | 0.085 | -0.117<br>(0.211) | 0.009 |
| Health spending | 0.117 | -0.226<br>(0.332) | 0.254 | 0.136 | -0.266<br>(0.374) | 0.211 |
| Social Mobility | 0.098 | 0.112<br>(0.240) | 0.754 | 0.094 | -0.114<br>(0.298) | 0.147 |
| Share of old population | 0.097 | -0.040<br>(0.235) | 0.392 | 0.208 | 0.311<br>(0.263) | 0.968 |
| Air Pollution | 0.091 | 0.021<br>(0.163) | 0.577 | 0.088 | 0.117<br>(0.223) | 0.979 |
| Population density | 0.090 | 0.054<br>(0.179) | 0.873 | 0.166 | -0.302<br>(0.255) | 0.009 |
| Share of young population | 0.088 | -0.039<br>(0.222) | 0.334 | 0.253 | -0.364<br>(0.238) | 0.003 |
| Quality of government | 0.082 | 0.019<br>(0.249) | 0.389 | 0.098 | -0.023<br>(0.347) | 0.422 |
| Commorbidities | 0.080 | -0.029<br>(0.169) | 0.279 | 0.091 | 0.094<br>(0.255) | 0.656 |
| Solar radiation | 0.079 | -0.039<br>(0.228) | 0.446 | 0.129 | -0.269<br>(0.321) | 0.144 |
| Infrastructure density | 0.078 | 0.019<br>(0.158) | 0.599 | 0.082 | -0.010<br>(0.258) | 0.406 |

Notes: The dependent variable in regressions is the total number of contagions per 100,000 inhabitants. Columns (1) to (3) reflect estimates of official contagions whereas Columns (4) to (6) those after correcting for under-reporting. The results reported correspond to the estimation of the top 100 models from all the possible regressions including any combination of the 15 variables. We use a fixed BRIC g-prior on the parameters and a Binomial model prior with prior mean model size equal to 7.5 in all cases. Variables are ranked by Column (1), the posterior inclusion probability. \* Significant at 10% level, \*\* significant at 5% level, \*\*\* significant at 1% level.

## Appendix B: Monte Carlo Experiment on Frequentist Model Averaging

This section presents the results of Monte Carlo experiments that investigate the performance of the Frequentist Model Averaging approach outlined in Section (6.2.2) when generating rankings of variable importance. In the Monte Carlo experiments, data on the potential explanatory variables in *Z* = [*ι_n_, z*_1_*, …, z*_9_] are generated from independent standard normal distributions. In the simulation study, *Z* is of dimension *n* × 10. We consider a sample size of *n* = 100. The dependent variable *y* is generated according to the following data-generating process:

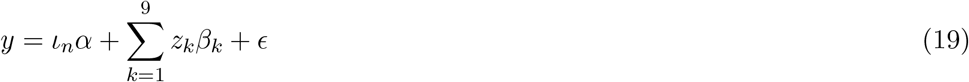

where 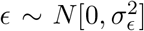. Notice that by adjusting *σ*^2^ it is possible to control the signal-to-noise ratio in the model. Values for *σ*^2^ are set to 1, 2, 5 and 10 in the different experiments, to asses the ability of the FMA approach to rank correctly the importance of *z_k_* ∈ *Z* under different scenarios of the *R*^2^.

The true slope coefficients *β_k_* capturing the effect of *z_k_* in the data-generating process given by equation (19) are generated in a way such that the constant term and the first two (nonconstant) covariates *z*_1_ and *z*_2_ are likely to be important regressors in the model. Specifically, the corresponding coefficients are drawn from a normal distribution with mean zero and a relatively large variance of 10. The parameters corresponding to *z*_3_, *z*_4_, *z*_5_ and *z*_6_ are similarly drawn from a normal distribution with mean zero. However, with a level of 0.1, the variance for these parameters is set to be low, implying that these covariates are less likely to be important. The variance of the parameters related to *z*_7_, *z*_8_ and *z*_9_ is set to 0.00001to reflect the variables have a negligible influence on *y*.

We focus on the Jackknife FMA approach based on different sizes of the selection matrix *S* of size *s × k* + 1 for different sizes of *s*. Due to the computational burden, we use *S* instead of estimating the 2^*k*+1^ models as in many empirical applications the enumeration of all the models might be quite time-consuming or infeasible. The design of *S* goes as follows. Each of the entries of *S* is filled by drawing from a Binomial distribution adjusted such that the expected probablity of *z_k_* being in the model is the 50% (i.e, by setting *φ* = 0.5 in *p* (*z_k_*) = *φ^k^* (1 − *φ*)^*K*−*k*^, which governs the probability of inclusion for each *z_k_* and *s*. This implies that on average, the number of regressors in each model is 5 (i.e, each row of *S_i,_*_∗_ has 5 ones and 5 zeros). As the total number of model combinations with *k* + 1 = 10 is 1024 and in applied analysis with large numbers of regressors, storage of the full model space may require unfeasible RAM memory, we just consider scenarios where *S* < 2^*k*+1^. In particular we consider S = 250, 500 and 1,000.

The results of the simulation exercise show are shown in Table 10. As observed, the group of top regressors conformed by the constant term, *z*_1_ and *z*_2_ always display weights above medium importance and low importance regressors, irrespective of the signal-to-noise ratio and the size of the selection matrix S. Their variable weights are in most of the cases the 99.9% or the 100%. Similarly, medium-relevance regressors appear above the low-importance ones in most of the scenarios, which implies a high level of accuracy. The variance of the residual does not appear to significantly affect the quality of the rankings based on FMA weights. However, the quality of the rankings appear to be dependent on the size of the sample of models S considered to derive the weights. This can be seen by looking at the quality of rankings produced with S = 1000 and S = 250. When the selection matrix S just considers a low number of models (i.e 250 models), a low-importance regressor *z*_8_ may obtain a variable weight within the range of medium-level of importance regressors *z*_3_ to *z*_6_. Despite this exception, we find that overall, the employment of FMA is accurate at ranking the groups of regressors by its importance.

**Table 10:** Monte Carlo FMA.

| S = 250 |  |  |  |  |
| --- | --- | --- | --- | --- |
| Variable | $R^2 = 0.977$<br>$\sigma^2 = 1$ | $R^2 = 0.949$<br>$\sigma^2 = 2$ | $R^2 = 0.822$<br>$\sigma^2 = 5$ | $R^2 = 0.628$<br>$\sigma^2 = 10$ |
| constant | 1.000 | 1.000 | 1.000 | 1.000 |
| $z_1$ | 0.9922 | 0.9711 | 0.9259 | 0.8591 |
| $z_2$ | 0.9934 | 0.9632 | 0.9201 | 0.8549 |
| $z_3$ | 0.6086 | 0.6893 | 0.7539 | 0.7670 |
| $z_4$ | 0.6637 | 0.5153 | 0.5466 | 0.5461 |
| $z_5$ | 0.5698 | 0.1234 | 0.0423 | 0.0255 |
| $z_6$ | 0.5536 | 0.1977 | 0.0958 | 0.0901 |
| $z_7$ | 0.0867 | 0.0151 | 0.0113 | 0.0144 |
| $z_8$ | 0.7158 | 0.6929 | 0.7275 | 0.7354 |
| $z_9$ | 0.0199 | 0.0049 | 0.0128 | 0.0237 |
| S = 500 |  |  |  |  |
| | $R^2 = 0.9752$<br>$\sigma^2 = 1$ | $R^2 = 0.9277$<br>$\sigma^2 = 2$ | $R^2 = 0.7810$<br>$\sigma^2 = 5$ | $R^2 = 0.534$<br>$\sigma^2 = 10$ |
| constant | 1.0000 | 1.0000 | 1.0000 | 1.0000 |
| $z_1$ | 0.9749 | 0.9574 | 0.8998 | 0.8093 |
| $z_2$ | 0.9798 | 0.9612 | 0.9022 | 0.8201 |
| $z_3$ | 0.4717 | 0.4762 | 0.5363 | 0.5595 |
| $z_4$ | 0.1060 | 0.0147 | 0.0021 | 0.0007 |
| $z_5$ | 0.4776 | 0.5130 | 0.5722 | 0.5888 |
| $z_6$ | 0.1570 | 0.0392 | 0.0065 | 0.0039 |
| $z_7$ | 0.0045 | 0.0009 | 0.0010 | 0.0013 |
| $z_8$ | 0.0140 | 0.0055 | 0.0077 | 0.0094 |
| $z_9$ | 0.0011 | 0.0010 | 0.0004 | 0.0001 |
| S = 1000 |  |  |  |  |
| | $R^2 = 0.978$<br>$\sigma^2 = 1$ | $R^2 = 0.939$<br>$\sigma^2 = 2$ | $R^2 = 0.794$<br>$\sigma^2 = 5$ | $R^2 = 0.579$<br>$\sigma^2 = 10$ |
| constant | 1 | 1.0000 | 1.0000 | 1.0000 |
| $z_1$ | 0.9825 | 0.9637 | 0.9136 | 0.8382 |
| $z_2$ | 0.9834 | 0.9698 | 0.9284 | 0.8680 |
| $z_3$ | 0.1681 | 0.0280 | 0.0006 | 0.0000 |
| $z_4$ | 0.3842 | 0.3448 | 0.3428 | 0.3570 |
| $z_5$ | 0.1352 | 0.0184 | 0.0014 | 0.0007 |
| $z_6$ | 0.1384 | 0.0155 | 0.0006 | 0.0001 |
| $z_7$ | 0.0041 | 0.0057 | 0.0085 | 0.0104 |
| $z_8$ | 0.0023 | 0.0028 | 0.0022 | 0.0012 |
| $z_9$ | 0.0014 | 0.0014 | 0.0003 | 0.0000 |

## Appendix C: Relative Importance Metric Computation

Let the variance of the dependent variable *Y* be given by 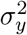, the variance of the set of regressors contained in *X* be denoted by Σ and the covariance of *Y* and the covariates by Σ_*yX*_. Let **P** denote the correlations among regressors and **P**_*yX*_ marginal correlations between regressors and *Y*, such that:

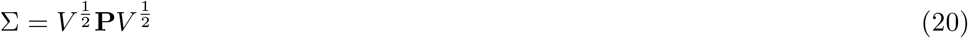

and

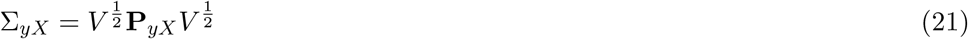

where *V* = *diag* (*V ar*(*X*_1_)*, …, V ar*(*X_p_*)). Defining the correlation between the model estimates and Y as 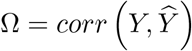, then the squared multiple correlation coefficient is expressed as:

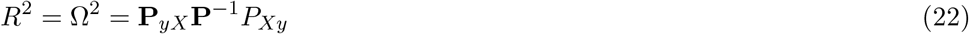

Then, the unexplained variance can be written as 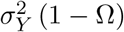 and the explained variance of a model with *X_k_* regresors with indices in the set *S* as 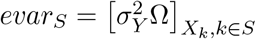. Finally, the sequential added explained variance when adding the regressors with indices in *M* to a model that already contains the regressors with indices in *S* as 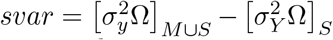.^21^ This implies that the true coefficient of determination is given by:

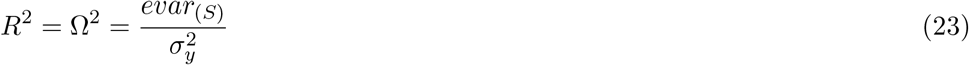

With these definitions in hand, for any model with *p* regressors, the R-squared can be expressed as:

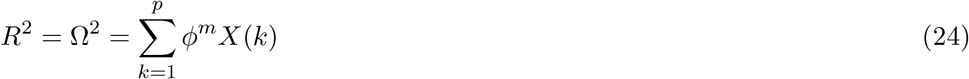

where *m* denotes the decomposition method.

The LMG method assigns to each regressor *X_k_* the following share:

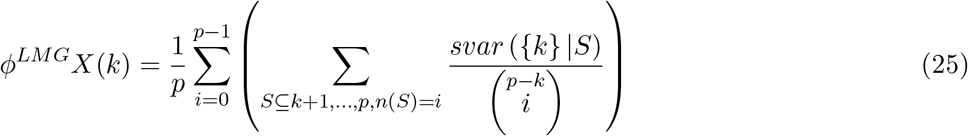

where *svar* denotes the sequentially added explained variance as defined above. Thus the share *φ_k_* assigned to regresor *k* is the average over model sizes *i* of average improvements in explained variance when adding regressor *k* to a model of size *i* that did not contain *k*. Hence, the LMG metric performs a *R*^2^ decomposition by averaging marginal contributions of independent variables over all orderings of variables and using sequential sums of squares from the linear model, the size of which depends on the order of the regressors in each particular model. On the other hand, the PMVD method assigns each regressor *X_k_* the following share:

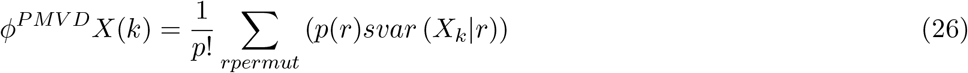

where where *p*(*r*) denotes the data-dependent pmvd weights. The idea is to use all the sequences *svar X_r,_k*+1 *, …, X_r_K* | (*X_r_*1 *, …, X_r_k*) for all *k* = 1*, …, K* − 1 to determine the weights of all orders *r* which are proportional to:

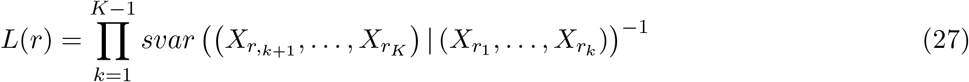

such that 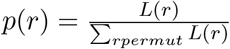.

Finally, to check the robustness of our results we also compute two alternative metrics of relative importance: (i) the Genizi (1993) and the (ii) CAR scores Zuber and Strimmer (2011)). The weights associated to the Genizi (1993) and CAR measures are given by:

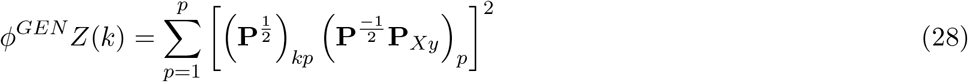

and

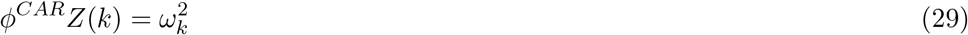

with 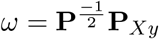.

## Footnotes

1 This methodological approach, has been used in economics to investigate the determinants of complex and multi-faceted processes in the field of economic growth (Sala-i-Martin *et al*., 2004, Crespo-Cuaresma *et al*., 2014), income inequality (Hortas-Rico and Rios, 2019), natural resources (Arin and Braunfels, 2018) or regional resilience (Rios and Gianmoena, 2020) among other topics.

2 An example of issues when registering and compiling data in a similarly developed and decentralized country is that of Spain. In Spain, in April 2, the Spanish Health Ministry published a note correcting their daily reports recognizing that some regions where counting differently hospitalizations to others, making impossible to compare the existing regional time series (SHM, 2020). Similar problems have occurred with the incidence data. Until April 15, active cases included PCR-confirmed cases for all regions, and serological tests for some regions. Starting on April 15, a change of methodology led to a split in both figures for some regions; by April 18 most had switched to the new methodology except for Galicia, which made the switch from April 28 onwards. Detailed press reviews on counting problems across regions on other variables such as the number of deaths in Spain are (Mouzo, 2020; Galaup and Sanchéz, 2020).

3 To that end, we correct the time series of reported cases using a delay-adjusted crude fatality ratio (CFR).

4 Indeed, in Italy there exists a multi-level governance structure of the regional health systems, such that major fiscal, financial and managerial responsibilities have been devolved to the regional level since the year 2001.

5 In any case, with the aim of verifying if this potential problem matters in the conclusions we follow Russell *et al*. (2020) and we use a delay-adjusted case fatality ratio to estimate under-reporting for each region and each date in Section 6.2.2.

6 The contiguous neighboring regions of Lombardy are Veneto, Trento, Piedmont and Emilia Romagna.

7 Information on the size and date of each regional peak and the cumulative cases by April 15 is provided in Table (A1) in the Appendix.

8 The corresponding regional NUTS-2 codings and regional names shown in the maps are: ITC1-Piedmont, ITC2-Aosta Valley, ITC3-Liguria, ITC4-Lombardy, ITF1-Abruzzo, ITF2-Molise, ITF3-Campania, ITF4-Apulia, ITF5-Basilicata, ITF6-Calabria, ITG1-Sicily, ITG2-Sardinia, ITH1-Autonomous Province of Bolzano, ITH2-Autonomous Provincce of Trento, ITH3-Veneto, ITH4-Friuli-Venezia Giulia, ITH5-Emilia-Romagna, ITI1-Toscana, ITI2-Umbria, ITI3-Marche, ITI4-Lazio

9 On the other hand, an insignificant and weak negative correlation coefficient of the –0.0891 arises between relative humidity and the magnitude of the regional pandemic (see Figure A1 of the Supplementary Material Appendix.

10 For a detailed discussion on the relevance of testing when monitoring the COVID-19 epidemic see Romer (2020), where it is argued that even a bad test can help guide the decision to isolate.

11 We consider 10 potential explanatory variables in our baseline analysis. Thus, the cardinality of the model space in this context is 2^10^ = 1024 models, based on different combinations of regressors. In Section (6.2.4) 15 regressors are employed, which implies a model space consisting of possible 32,768 models.

12 In particular, the g-prior hyper-parameter takes the value of 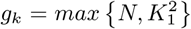 such that *g*(*η*) ∼ 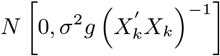. The Binomial prior on the model space, regulates prior model probabilities according to *p* (*M_k_*) = *φ^k^* (1 − *φ*)^*K*1−*k*^, where each covariate *k* is included in the model with a probability of success *φ*. We set *φ* = 0.5 which implies a prior model size of 5 regressors.

13 A common concern in BMA is that PIPs may simply reflect the strength of the correlations between the explanatory variables and the dependent variable. However, this is not an issue here since the correlation between the PIPs and the pairwise correlation between contagions and the set of potential drivers, in this context is about 0.43. This suggests the probabilistic importance ranking obtained from the analysis performed here contains valuable information that cannot be learned from the mere computation of correlations.

14 We do not resort to conventional Monte Carlo Markov Chain Model Composition (*MC*^3^) algorithms as the enumeration of all the models is computationally feasible in this context.

15 For an alternative graphical representation of the results in Table (4) see Figure (A2) in the Appendix.

16 We do not attempt to estimate both symptomatic and asymptomatic cases as it is not clear yet what is the precise share of asymptomatic and in the context of the approach set out by Russell *et al*. (2020) this adjustment would just change the estimation by a scalar *k* leaving the ordering and the variability unchanged.

17 We assumed the delay from confirmation-to-death followed the same distribution as estimated hospitalisation-to-death, based on data from the COVID-19 outbreak in Wuhan, China, between the 17th December 2019 and the 22th January 2020. Thus, the distribution used is a Lognormal with a mean delay of 13 days and a standard deviation of 12.7 days.

18 We also provide the corresponding estimates of the BMA robustness checks shown in Tables (7) and (8) when using the corrected contagion series in the Tables (A3) and (A4) of the Appendix A.

19 The EQI is built upon three different pillars that refer to the degree of impartiality, corruption and quality of public services and it is based on survey data about the perceptions and experiences of European citizens on the quality, impartiality and level of corruption in education, public health care and law enforcement.

20 Figure A3 in the Supplementary Material Appendix displays the cumulative model probability plots providing evidence of a strong concentration of probability density in the top models 100. The top 100 highest probability models, cover approximately the 10% of the model space and concentrate the 89.76% of the posterior probability mass.

21 For a given regressor, if we let *S_k_*(*r*) denote the set of regressors entered into the model before regressor *X_k_* in the order *r*, the portion of *R*^2^ allocated to regressor *X_k_* in the order *r* is given by *svar* (*X_k_*|*S_k_*(*r*)) = *R*^2^ (*X_k_* ∪ *S_k_*(*r*)) − *R*^2^ (*S_k_*(*r*)).

